# Explainable fNIRS Based Pain Decoding Under Pharmacological Conditions via Deep Transfer Learning Approach

**DOI:** 10.1101/2023.11.15.23298553

**Authors:** Aykut Eken, Sinem Burcu Erdoğan, Gülnaz Yükselen, Murat Yüce

**Author notes:** Corresponding author Aykut Eken, TOBB University of Economics and Technology, Söğütözü, Söğütözü Cd. No:43, 06510 Çankaya / Ankara / Turkey. The authors equally contributed to writing of the manuscript (Aykut Eken, Sinem Burcu Erdoğan).

## Abstract

**Problem Statement:** Pain has a crucial function in the human body acting as an early warning signal to protect against tissue damage. However, both assessment of pain experience and its clinical diagnosis rely on highly subjective methods. Objective evaluation of the presence of pain under analgesic drug administrations becomes even more complicated.

**Objectives:** The aim of this study was to propose a transfer learning (TL) based deep learning (DL) methodology for accurate detection and objective classification of the neural processing of painful and non-painful stimuli that were presented under different levels of analgesia.

**Method:** A publicly available fNIRS dataset of 14 participants was obtained during an experimental protocol that involved painful and non-painful events. Separate fNIRS scans were taken under the same nociceptive protocol before analgesic drug (Morphine and Placebo) administration and at three different times (30,60 and 90 min) post-administration. By utilizing data from pre-drug fNIRS scans, a DL architecture for classifying painful and non-painful stimuli was constructed as a base model. Knowledge generated in pre-drug base model was transferred to 6 distinct post-drug conditions by adapting a TL approach. The DeepSHAP method was utilized to unveil the contribution weights of nine R OIs for each of the pre-drug and post-drug models.

**Results:** Mean performance of pre-drug base model was above 95% for accuracy, sensitivity, specificity and AUC metrics. Each of the post-drug models had mean accuracy, sensitivity, specificity and AUC performance above 90%. No statistically significant difference across post-drug models were found for classification performance of any of the performance metrics. Post-placebo models had higher decoding accuracy than post-morphine models.

**Conclusion:** Knowledge obtained from a pre-drug base model could be successfully utilized to build pain decoding models for six distinct brain states that were altered with either analgesic or placebo intervention. Contribution of different cortical regions to classification performance varied across the post-drug models.

**Importance:** The proposed methodology may remove the necessity to build new DL models for data collected at clinical or daily life conditions for which obtaining training data is not practical or building a new decoding model will have a computational cost. Unveiling the explanation power of different cortical regions may aid the design of more computationally efficient fNIRS based BCI system designs that target other application areas.

## 1. Introduction

Pain is a vital function of the human body which serves as an early warning signal to protect tissue damage. The extent to which an individual experiences pain still remains a complex and subjective phenomenon, and is considered to depend on a variety of intrinsic and extrinsic factors that include the efficiency of communication between the nociceptors and their subcortical and cortical projections (De Felice & Ossipov, 2016) besides genetics, past experiences and cultural influences. While the current common methodology for assessing perception of pain and its intensity level relies on self-reports in clinical practice, there may be conditions where patients are unable to provide verbal self-reports such as a surgical procedure performed under anesthesia or in situations where the patient is unconscious due to a variety of conditions such as critical cerebral tissue damage. Patients with severe cognitive impairments or patients who preserve their mental abilities but who are unable to communicate with their external environment may also be unable to provide objective and accurate self-reports of their pain experience.

Pain has few biomarkers that can be used in clinical practice (Woo et al., 2017). Some biomarkers are intended to track pain intensity and complement self-reports as a way of assessing the incidence or intensity of pain, while others are intended to reveal underlying pathobiological conditions (Woo et al., 2017). However, in the above mentioned situations, there is a lack of an objective biomarker of pain that can aid precise evaluation and management of treatment procedures. Objective evaluation of pain perception would have numerous clinical advantages, including the ability to continuously monitor and assess neural correlates of perceived pain intensity during surgery and quantitative evaluation of the progress and efficacy of a treatment strategy. Such an objective evaluation marker could assist execution of operational procedures under optimal conditions through adjustment of the analgesic regime when required.

Previous functional neuroimaging studies conducted with positron emission tomography (PET), functional magnetic resonance imaging (fMRI) and functional near infrared spectroscopy (fNIRS) demonstrated consistent pain related localized hemodynamic responses in the human brain (Morton et al., 2016; Paquette et al., 2018; Yucel et al., 2015). Moreover, these studies also demonstrated spatial and temporal differences in the neural processing of low and high intensity painful stimuli (Bornhovd et al., 2002; Morton et al., 2016; Paquette et al., 2018; Yucel et al., 2015). Pain induced deactivation in the medial prefrontal cortex (mPFC) regions during both acute and chronic conditions has also been consistently observed across different neuroimaging studies conducted with different modalities (Karunakaran et al., 2021; Ong et al., 2019; Ozturk et al., 2021). Moreover, morphine induced attenuation of the deactivation in the MPFC during painful stimuli processing was also reported in the study of Peng et al (2018). Overall, these studies have provided valuable insights into the neural mechanisms underlying pain processing in the human brain. Besides, they also addressed the promise of exploring robust biomarkers of pain processing under analgesic or different drug administrations.

Among these modalities, fNIRS has shown a great potential for extracting objective biomarkers of pain in the clinical or operative environments due to its numerous advantages such as ability to collect hemodynamic data noninvasively with wearable ergonomic probes that can be placed at the surface of the scalp. Previous studies using fNIRS have consistently demonstrated significant changes in oxygenated hemoglobin (HbO) concentration in the prefrontal cortex (PFC) in response to painful stimuli, including cutaneous (Green et al., 2022), dental (Racek et al., 2015) and visceral pain (Becerra et al., 2016). These observations are supported by findings from fMRI studies reporting deactivations in anterior PFC blood oxygenation level-dependent (BOLD) signals following painful stimuli (Gundel et al., 2008; Kong et al., 2010; Lui et al., 2008; Tseng et al., 2010). Under analgesic state, Beccera et al (2016) obtained hemodynamic recordings from the PFC during a colonoscopy procedure (Becerra et al., 2016). Analysis of fNIRS data revealed a specific, reproducible PFC activity corresponding to the time intervals when patients grimaced. The pattern of activation was similar to that obtained in previous studies in awake healthy individuals while they were exposed to nociceptive stimuli. Similar hemodynamic activation patterns obtained during painful events under both awake and sedative conditions suggest that unsuccessful inhibition of the neuronal processing of a nociceptive stimulus due to insufficient levels of analgesia can be objectively quantified with fNIRS derived biomarkers. Karunakaran et al (2023) also showed that the use of fNIRS during knee surgery can provide objective measures of pain-related brain activity (Karunakaran et al., 2023). After analysing fNIRS data obtained during pre, intra and postoperative stages, they found a decrease in resting-state functional connectivity (FC) within the mPFC during the postoperative state when compared to the preoperative awake state. Also, they observed that negative intraoperative FC between the mPFC and somatosensory cortex (S1) was associated with higher reported postoperative pain levels. As a conclusion from this study, it can be inferred that neurophysiological information obtained from fNIRS recordings during surgery can provide objective measures of pain-related brain activity. In a study by Kussman et al. (2016) involving patients undergoing catheter ablation of arrhythmias, somatosensory and frontal cortical hemodynamic activations were measured with fNIRS (Kussman et al., 2016). The results showed that the frontal cortical signals were suitable for analysis and a decrease in HbO concentration in response to the ablative lesions was observed. These cortical signals mirrored the responses seen in awake, healthy volunteers and findings from other studies involving nociceptive stimulation. These studies highlight the feasibility and potential utility of fNIRS as an objective measure of cortical activation during nociceptive procedures under general anesthesia.

Despite the promising results, there are challenges in using fNIRS-derived neural markers for accurate detection of pain. One issue is the presence of habituation effect which results in a decrease in the amplitude of hemodynamic responses to repeated painful stimuli over time (Yucel et al., 2015). In addition, the shape of the hemodynamic response function (HRF) obtained during painful stimuli present intra and inter subject variability which has also been shown to be dependent on cortical regions and stimuli types (Yucel et al., 2015). One major limitation for deriving robust pain biomarkers from both fNIRS or fMRI signals via mass univariate statistical approaches relies on the low spatial and functional sensitivity of these techniques since the achieved spatial resolution spans millions of neurons with diverse functional properties and distributed connections across different layers. Another issue that has been noted in the use of fNIRS for detection of pain is the effect of analgesics, specifically opioids such as morphine, on the hemodynamic response. Peng et. al. (2018) found that morphine administration was associated with an attenuated HbO signal in the medial portion of the anterior prefrontal cortex (Brodmann Area 10) in response to painful stimuli (Peng, Yucel, et al., 2018).

Evaluation of hemodynamic and behavioral correlates of different levels of pain intensity is performed by use of conventional statistical approaches which provides an insight at the population level and does not allow inferences to be made at the single subject or single stimulus level. Due to these limitations, accurate detection and objective identification of perceived pain intensity under different pharmacological conditions and how this perception evolves over time is a challenging problem. Within this context, deep learning (DL) techniques may provide a more effective approach to the problem of decoding intensity level of perceived pain from neural processing information of a nociceptive stimulus obtained with functional neuroimaging modalities. DL methodologies provide several benefits such as integration of all available biological data into a single ‘best prediction’ about the output of the algorithm besides their ability to capture information across multiple spatial scales.

Several studies implemented DL methods to fMRI and fNIRS signals to search for “fingerprints” specific to acute pain processing. Rojas et al (2021) aimed to develop an objective tool for assessing pain in non-verbal patients using DL models and fNIRS data (Rojas et al., 2021). The authors explored the utility of different DL models and compared their performance in accurate identification of pain. The study found that combination of forward and backward information in the Bidirectional Long Short-Term Memory (Bi-LSTM) model achieved a 90.6% accuracy in two class classification of pain intensity level. The use of DL models eliminated the need for complex feature extraction procedures and reduced subjectivity in designing hand-crafted features when compared to supervised machine learning models. These findings represented a step forward in the development of a physiologically-based diagnosis of perceived pain intensity and can assist clinicians in objective assessment of pain in non-verbal patients.

In order to overcome the above mentioned challenges associated with decoding the presence of neural processing of nociceptive stimuli and improve the sensitivity of fNIRS recordings to identification of the presence of nociceptive stimuli processing at the single stimulus level while the subject is under different analgesic conditions, the aim of this study is to propose a transfer learning (TL) based DL methodology for accurate detection and objective classification of painful and non-painful stimuli that are presented under different levels of analgesia. TL is a specific supervised learning method which involves transfer of knowledge (i.e., feature weights) from a pre-trained base model to a new model that is utilized to make inferences about a similar population data after addition of a few computationally efficient fine-tuning steps (Wu et al., 2022). Within the context of proposed work, the TL approach was utilized to transfer knowledge (i.e., weights) of the constructed DL model from pre-drug fNIRS scans and the base neural network knowledge of the pre-drug DL model was adapted to the problem of two class classification of the neural processing of two levels of painful stimuli collected under two different pharmacological interventions and at three time points post-intervention (i.e., 30 min, 60 min, 90 min)(Peng, Yucel, et al., 2018).

To date, there have been no studies that have shown the efficiency of TL for single trial classification of the presence of painful stimuli processing under different pharmacological conditions. This approach might be prominent for two potential applications: 1) we demonstrate the potential of training DL models for specific classification problems where a baseline fNIRS data is available and 2) a model trained with this baseline data can be adapted to data collected at different clinical or daily life conditions where obtaining training data is not feasible/practical to build novel ML or DL models.

The presented work addresses two main research questions. Our first question aimed to assess the feasibility of implementing a TL methodology to decode the neural processing of two levels of nociceptive stimuli obtained under two distinct pharmacological interventions and at different times post-intervention. Our hypothesis was to test whether we can decode the neural processing of low and high-level painful stimuli states by utilizing hemodynamic responses obtained before and after a morphine or a placebo drug administration with well defined performance parameters such as area under the curve (AUC) being greater than 0.9 which is accepted as an excellent classification performance (0.9-1) according to previous clinical studies (Han, 2022; Mandrekar, 2010; Metz, 1978). Our second research question aimed to assess the contribution of features obtained from different cortical regions to the classification performance of the proposed DL model and how this contribution changes as hemodynamic activity is modified with morphine or placebo intervention. For this purpose, an explainable artificial intelligence (xAI) method named DeepSHAP which combines Shapley values obtained from Shapley Additive Explaination Method (Lundberg & Lee, 2017) with the DeepLIFT algorithm (Shrikumar et al., 2017) was utilized. Unveiling the explanation power of different regions of interest is prominent as it may aid the design of more computationally efficient brain computer interface (BCI) system designs that target pain detection and such an approach may provide more precisely localized physiological markers of pain.

## 2. Materials and Methods

### 2.1. Dataset

An fNIRS dataset that was previously published in (Peng, Yucel, et al., 2018) was utilized in the presented work. In this study, 14 male volunteers who had no recent history of pain or opioid abuse were recruited. Each subject had two site visits where he was administered with either an oral morphine or a placebo pill. At each site visit, fNIRS scans were taken during a nociceptive stimuli protocol a) before and b) after administration of an oral morphine or a placebo pill. The pills looked identical and the order of placebo or morphine administration was randomized.

At each site visit, the subject had an fNIRS scan prior to drug administration during a nociceptive stimuli protocol which consisted of 6 painful and 6 non-painful stimuli that were delivered to the left thumb with an electrical stimulator. Criteria for determining the electrical threshold for low level pain and high level pain conditions were explained extensively in the study carried out by Peng and colleagues (Peng, Yucel, et al., 2018). Each nociceptive stimulus lasted for 5 seconds followed by a 25 second rest period. The same nociceptive stimuli paradigm was applied to participants at separate fNIRS sessions that took place after 30 min (Post-Morphine-30 (PM-30)), 60 min (Post-Morphine-60 (PM-60)), and 90 min (Post-Morphine-90 (PM-90)) of morphine administration and after 30 min (Post-Placebo-30 (PP-30)), 60 min (Post-Placebo-60 (PP-60)) and 90 min (Post-Placebo-90 (PP-90)) of placebo administration. fNIRS recordings were collected from the medial portion of the frontopolar cortex (FP, medial Brodmann Area 10), the right primary S1 and a portion of the left lateral PFC.

### 2.2. Regional Information

The publicly available fNIRS dataset included real head coordinates of source and detector positions for each subject and scan. These real head coordinates were converted to MNI coordinates through the Colin 27 atlas (Holmes et al., 1998) by use of NIRS-SPM toolbox (Ye et al., 2009) to reveal the corresponding cortical region. The head coordinates of individual optodes and channels were extracted for each drug administration scan of each subject. After estimation of the MNI coordinates from real head coordinates, the MNI coordinates of pre-scan session of morphine and placebo administration of 14 subjects were averaged for each optode position. Table 1 demonstrates the mean MNI coordinates of each channel and the relevant standard deviation across subjects averaged across all scans (Okamoto et al., 2004). After spatial registration of optode coordinates to the MNI space, 10 cortical regions were determined which included the right primary motor cortex (R MI), right somatosensory cortex (R SI), right and left pre motor cortices (R & L PMC), left inferior frontal gyrus (L IFG), right and left frontopolar area (R & L FPA), right and left dorsolateral prefrontal cortices (R & L DLPFC) and right supramarginal gyrus (R SMG). For morphine administration scans, the 10 cortical regions of interest were determined as R MI, R SI, R & L PMC, L IFG, R & L FPA, R & L DLPFC and R SMG. For placebo session, real coordinates corresponded to 9 cortical regions of interest that included R SI, L the IFG, R & L PMC, R & L FPA, R & L DLPFC and R SMG.

### 2.3. Dataset Preparation

#### 2.3.1. Data Preprocessing and Trial Extraction

fNIRS data preprocessing was performed with HomER3 toolbox (Huppert et al., 2009). Light intensity data were first converted to optical density (OD) by use of the Beer-Lambert law formula 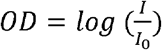 where *I*_0_ represents the intensity of incident light that is emitted through a light source placed at the surface of the scalp tissue and *I* represents the intensity of light collected at the channel forming detector at each time point. Motion artifacts were removed from OD data by a hybrid approach where Wavelet transform (Molavi & Dumont, 2012) and PCA (Zhang et al., 2005) were applied consecutively in order to preserve as many trials as possible unlike the process followed in the original study. After motion artifact removal, a Butterworth band-pass filter with high and low cut-off frequencies of 0.01 and 0.1 Hz were applied to remove heart beat (>1 Hz), respiration (0.15-0.4 Hz) (Fekete et al., 2011) and Mayer waves (∼0.1 Hz) (Yucel et al., 2016). Oxyhemoglobin concentration changes (Δ*HbO*) and deoxyhemoglobin concentration changes (Δ*Hb*) were estimated by using the Modified Beer-Lambert law (Cope et al., 1988). For each channel Δ*HbO* data, a general linear model (GLM) based short-channel regression was applied to remove the global systemic noise where the global systemic noise was modeled with the preprocessed Δ*HbO* signal of the closest short channel. Let *S* and *L* represent the time series Δ*HbO* data at short and long channels consequently. To perform the regression of systemic noise from long channels, the beta coefficients of short channels (*β*_*short*_) were estimated by using the equation below;

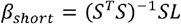

The scaling coefficient *β*_*short*_ was used to linearly regress out the systemic noise recorded by short-channels by using equation below;

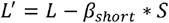

For each trial, a pre-stimulus period of 1 second and a 30 second period after onset of each stimulus (i.e., 5 seconds of electrical stimulus application and 25 seconds of resting period) were truncated. Each trial block was down sampled to 1 Hz in order to reduce the computational complexity during model training. For each subject, this data were organized in a matrix form with dimensions set as *number of trials (N)x number of time points (T) x number of channels* (C) for *“Painful”* or *“Non-painful”* stimuli classes.

For each experimental session, the time series data represented as D were reorganized as *D* = {(*X*_1_,*Y*_1_), (*X*_2_,*Y*_2_),,… … .., (*X*_*N*_*Y*_*N*_)} where X_i_ is a 3 dimensional matrix with dimensions of *number of trials x number of time points x number of channels* for each subject i and *Y* represents the corresponding stimulus intensity (i.e., painful or non-painful *Y* = {−1,1}) for each element of Xi. Since 2 pre drug sessions existed for each subject, 336 labelled trials (i.e., 2 sessions x 14 subjects x 6 trials x 2 intensity levels) were obtained from the pre drug sessions and 168 labelled trials (i.e.,14 subjects x 6 trials x 2 intensity levels) were obtained from each post drug session (i.e., *PM-30, PM-60, PM-90, PP-30, PP-60* and *PP-90*). Hence, the feature matrix had dimensions of *336 x 31 x 24* for pre-drug session and *168 x 31 x 24* for each post-drug session.

### 2.4. Deep Learning Steps

After preprocessing and reorganization of the fNIRS time series data, the DL model training steps included i) data augmentation, ii) implementation of the deep neural network (DNN) architecture design and iii) adapting the TL approach to post-drug datasets. During DNN training, only *ΔHbO* data were utilized due to higher SNR compared to *ΔHb* (Montero-Hernandez et al., 2018). Tensorflow toolkit (version 2.8.0) (Abadi et al., 2016) was utilized to construct and design the DNN architecture and for further application of the TL approach to each of the six post-drug data set. This procedure was repeated 30 times by randomizing the data augmentation step (the details of this step is explained at section 2.4.1) and averaging all loss and accuracy results. Pipeline depicting the order of analysis steps is shown in Figure 1.

**Figure 1:**
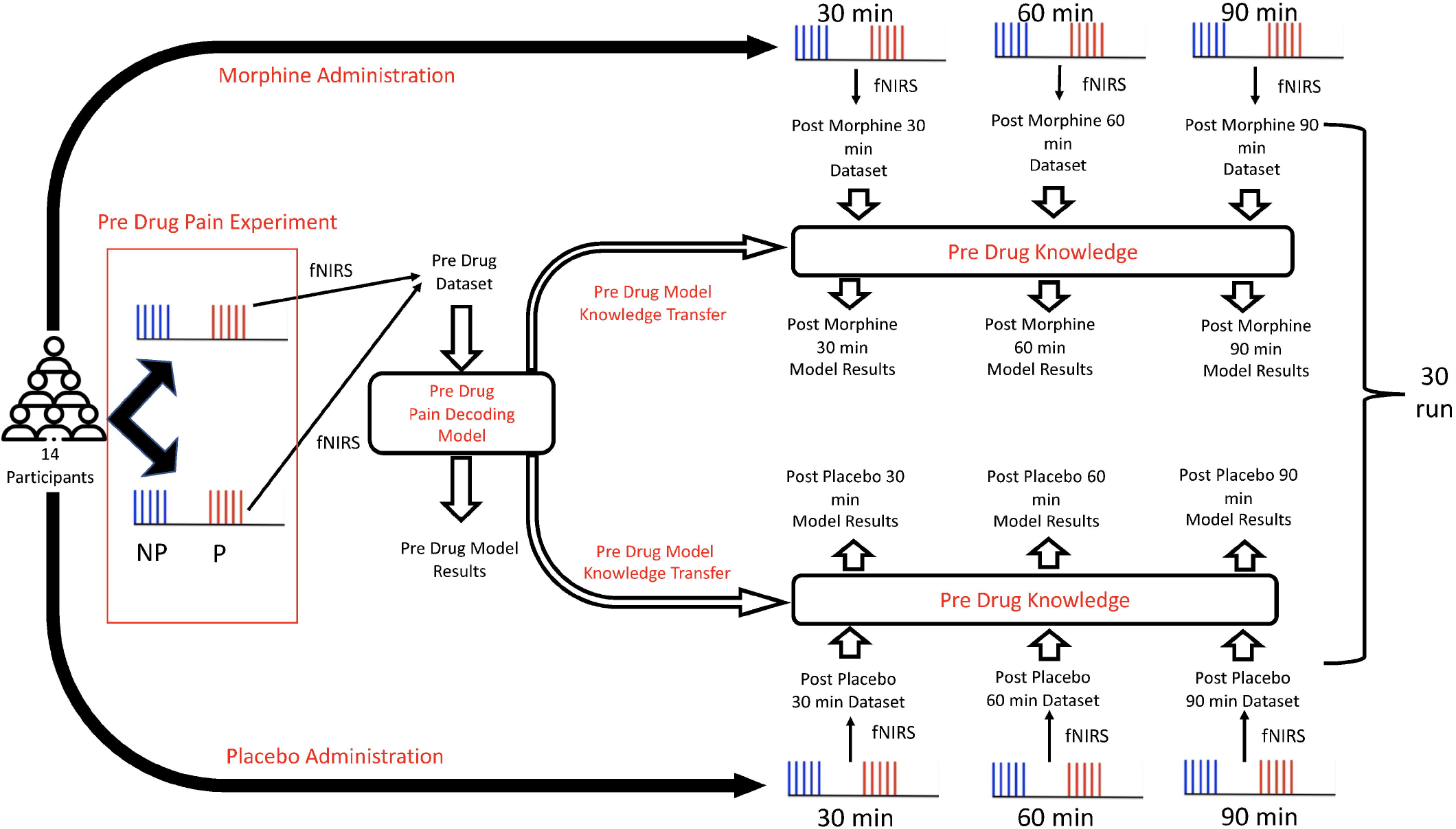
Pipeline of the analysis steps. P: Pain, NP: Non-pain.

#### 2.4.1. Data Augmentation

The pre-drug dataset for constructing the baseline model was split into %60 training, %20 test and %20 validation sets. The “train_test_split()” function of scikit-learn toolkit (Pedregosa et al., 2011) was utilized to perform this operation. At each run, training, test and validation data sets were randomized. Data augmentation was performed due to relatively small sample size of the available fNIRS data when compared to other application areas of deep learning (e.g., automation, finance). This step was applied only to the training data set. For the data augmentation procedure, time-domain approaches were applied to each truncated *ΔHbO* trial time series which involved either addition of a linear trend or Gaussian noise (Wen et al., 2021). The linear trend addition procedure involved addition of linear trends whose slope values were randomly chosen as 0.01, 0.05 or 0.1. These slope values were randomly selected and added to truncated *ΔHbO* time series of each trial of each channel. The second approach involved addition of Gaussian noise with zero mean and randomly selected variance (0.01, 0.05 and 0.1) to each trial time series data of each channel. After pooling single trial *ΔHbO* data from all channels and subjects *(i*.*e, 336 trial data (2 session*14 subjects*6 trial *2 stimulus intensity levels) x 31 time points x 24 channels)*, the training portion of this data set was augmented 25 times with randomized application of either of the above mentioned time-domain procedures.

#### 2.4.2. Proposed DNN Architecture

A DNN based on one-dimensional (1-D) convolutional layers was developed. In this network, three 1-D convolutional layers existed whose filter counts were 32, 64 and 128 with a convolution length of 2. Rectified linear unit (ReLU) layers were added as the activation function to the output of these layers. 1-D max-pooling layers with a length of 2 were added to the output of these ReLU layers. A dropout layer with a rate of 0.4 was added after every max pooling layer to avoid overfitting. After the third dropout layer, a flatten layer was added to convert the output of the final dropout layer as a single dimensional vector rather than a two dimensional one. The flattening layer was followed by addition of a dense layer with 256 units, a ReLU activation function and an additional dropout layer with a rate of 0.4. The final output layer consisted of the classification layer with a sigmoid function. Graphical representation of the designed network and its summary from Tensorflow are shown in Figure 2.

**Figure 2:**
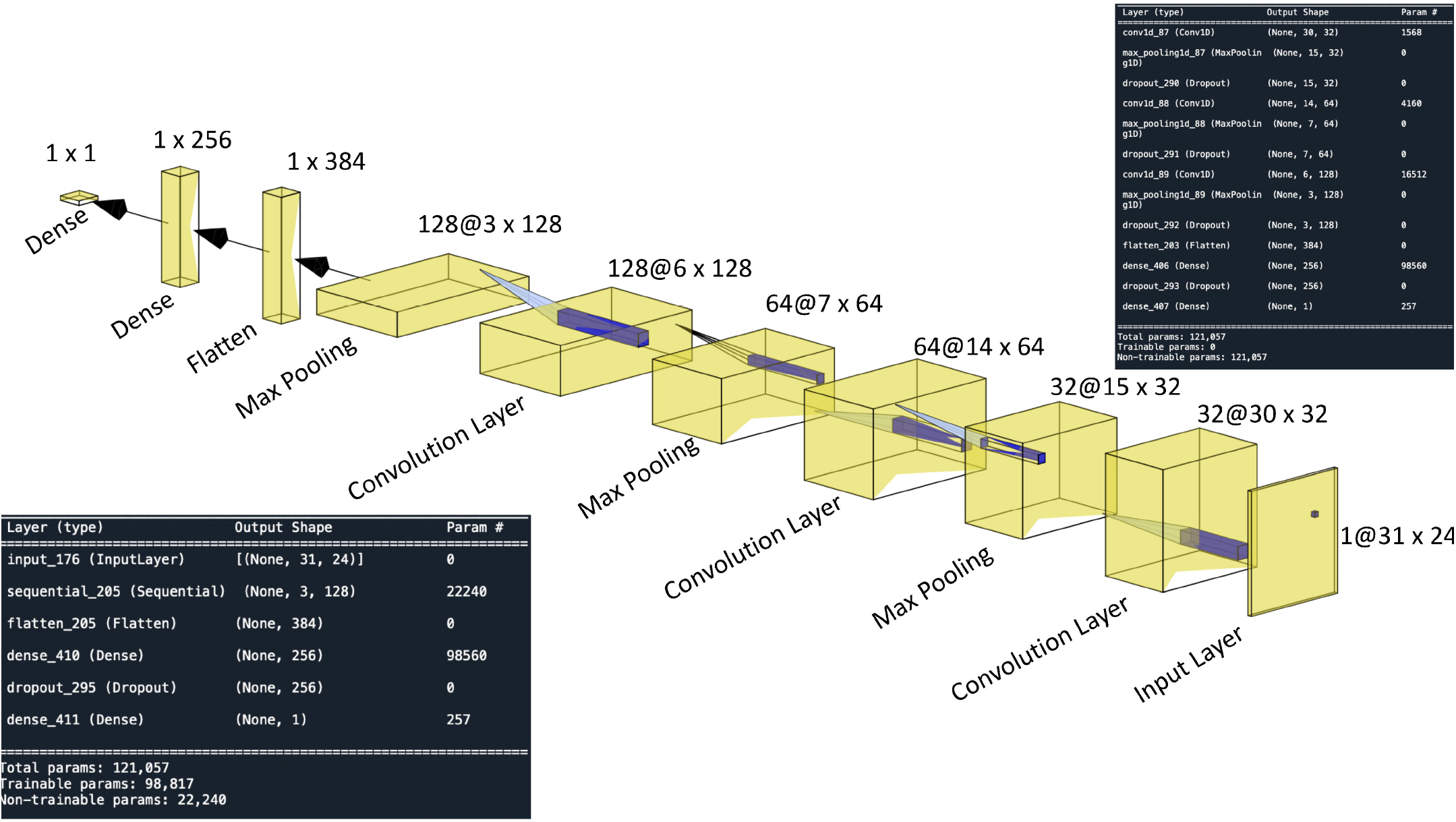
Graphical representation of the proposed DNN structure. Summary of the model architecture post TL procedure is given at the bottom left section. Numbers given on every layer indicate Filter size of layer @ Output size of layer.

Training the pre-drug model (PDM) involved use of Adam optimizer with a learning rate of *η* = 10^−4^. During each training session, a dynamic learning strategy was applied where the learning rate was reduced with a factor of 0.01 if validation loss did not change during 10 consecutive epochs and this strategy continued till the minimum *η* became 10^−6^. The batch size was 16 and the number of epochs was 100. For the post drug models (i.e., PM and PP models), training of added layers was carried out by using Adam optimizer with a learning rate of *η* = 10^−4^ and similar to the PDM training, the learning rate was reduced with a factor of 0.01 if validation loss did change during 10 consecutive epochs and/or till a minimum *η* of 10^−6^ was obtained. Batch size for post drug models was also 16 and the number of epochs was 100.

#### 2.4.3. Fine-Tuned Transfer Learning (TL) Approach

TL is a relatively new approach for developing neural decoding models (Peterson et al., 2021) especially for BCI applications (Azab et al., 2018). TL is based on the premise that knowledge generated from a pre-trained base model can be used to solve another similar classification problem on a novel data set (Wu et al., 2022). The PDM was constructed with fNIRS HbO data recorded during pre-drug sessions. Our purpose was to transfer knowledge generated from this pre-model to construct post-drug models obtained under two different pharmacological conditions (i.e., PM, PP) and at three time points post-intervention (30 min, 60 min, 90 min). Similar to the PDM, the post drug models took fNIRS signals collected during the same nociceptive paradigm as input. The rationale behind utilizing a TL approach relies on the assumption that such an adaptive training methodology would be able to capture the common neural signature of a nociceptive stimulation task obtained during dynamic brain states which were altered by a pharmacological intervention and this alteration would be expected to change with respect to time.

After training the PDM, the attained weights (i.e., knowledge) obtained in between the pre-trained layers that begun from the first convolutional layer to the last max-pooling layer were transferred to construct post drug models. For fine-tuning purposes, an additional flatten layer, a dense layer with 256 units, a dropout layer with a rate of 0.4 and a final classification layer with a sigmoid activation function were adjusted for each of the post-drug decoding networks separately. The final feature information was utilized to predict the label of stimuli (painful/non-painful) obtained during post morphine and post-placebo fNIRS scans of 30, 60 and 90 min post-drug administration sessions. The accuracy, sensitivity and specificity results are reported as an average of 30 runs. The painful events were labelled as positive (+) class and non-painful events were labelled as negative (-) class.

### 2.5. DeepSHAP Explanation

The DeepSHAP (Deep SHapley Additive exPlanations) method (Lundberg & Lee, 2017) was adapted to each model in order to evaluate the contribution of different cortical regions to model specific decoding performance. The SHAP approach is based on estimating a parameter named Shapley Value. Originally coming from cooperative game theory (Shapley, 1953), this value basically estimates the relative contribution of a feature to an output when compared to all possible other feature combinations. The DeepSHAP approach is defined as integration of SHAP method to the DeepLIFT algorithm in order to understand the feature specific contribution to the final classification decision (Shrikumar et al., 2017). The output of a neural network is decomposed to each input by performing backpropagation of neuronal contributions to every feature and SHAP values are estimated based on independence assumption of input features and linearity of the model.

For estimating Shapley values, attribution *ϕ*_*v*_ (*i*) for a feature *i* is obtained with the below formula:

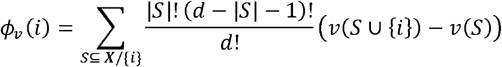

where ***X*** is the feature set that contains *d* number of features, *x* is the vector of feature values, represents the subset of features, *v* is the value function that takes *S* as input and *v*(*S*) is the prediction of the total contribution of the feature set 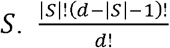 is the normalization term for subset *S* and *v* (*S* ⋃ {*i*}) − *v* (*S*) corresponds to feature *i*’s marginal contribution with respect to subset of features *S* and is averaged for *S* ⊆ ***X***/{*i*}. If function 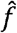 is considered as a model prediction function, *P*_*x∉s*_ is the probability of feature values that are not in the subset *S* and 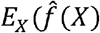 is the average predicted value, *v*(*S*) can be computed by using the below formula;

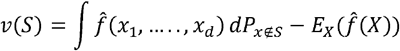

Total contribution of (*S*) has to satisfy four properties;

- Efficiency: Contributions of all features should be added up to difference (*v* (*S* ⋃ {*i*}) − *v* (*S*))) : ∑_*i*∉*X*_ *ϕ*_*v*_ (*i*) = *v*(*X*).
- Symmetry: If (*v* (*S* ⋃ {*i*}) = *v* (*S*) ⋃ {*j*}) for all *S* ⊆ ***X***/{*i,j*}, then *ϕ*_*v*_ (*i*) = *ϕ*_*v*_ (*j*)
- Dummy: If (*v* (*S* ⋃ {*i*}) = *v* (*S*) for all *S* ⊆ ***X***, then all *ϕ*_*v*_ (*i*) = 0, which means the feature does not have any contribution to output.
- Additivity: If there are two outcomes for a single case, *v*_1_ and *v*_2_, Shapley values have an additivity feature which can be represented as 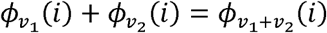.

Within the context of the proposed work, contribution of each input feature *(feature set: # of channels * # of time points per trial*) to the final decision of the network was computed for every run and the contribution of features extracted from all channels within each defined ROI (Table 1) was defined as the Shapley contribution of the relevant ROI. Therefore, a Shapley value matrix with dimensions of number of runs*number of ROIs was computed for each of the PDM and post drug models. After every model training, Shapley values were estimated by using DeepSHAP. For DeepSHAP explainer of PDM, test data had a size of 67 (# of trials) x 31 (# of time points) x 24 (# of channels). At each run, 67 test samples included data from both classes. Shapley values across all channels within each ROI were averaged in order to interpret the independent contribution of each ROI to classification performance. For post-drug sessions, the corresponding test data for estimating Shapley values had a size of 31 (# of trials) x 31 (# of time points) x 24 (# of channels).

### 2.6. Statistical Analysis

The accuracy, sensitivity and specificity performances of PDM and post-drug models were compared based on values obtained from 30 runs. For each performance metric, normality of performance results from all models were tested by using Shapiro-Wilk test. Because the distribution of values belonging to each of the performance metrics violated the normality assumption, the statistical comparison between PDM and post-drug model performances were carried out by using the Kruskal-Wallis test for accuracy, sensitivity and specificity metrics. Post-hoc comparisons were conducted with Bonferroni.

The classification performance of post-drug models were compared by using a 2 x 3 ([Morphine, Placebo] x [30 min, 60 min, 90 min]) repeated measures analysis of variance (ANOVA) after performing a box-cox transformation on all results due to non-normal distribution of data samples belonging to each of the performance metric.

## 3. Results

### 3.1. Deep Transfer Learning Model Performances

Training and validation accuracy curves of PDM are shown in Figure 3. The final training and validation accuracy scores of 30 runs reached to 0.99 ± 0.003 and 0.97 ± 0.02 for PDM. Training and validation accuracy curves of post drug models are shown in Figure 4. The training accuracy values reached to 1.0 after 10 to 15 epochs for the post drug models. For PP models, validation accuracies of PP-30, PP-60 and PP-90 reached to 0.91 ± 0.05, 0.90 ± 0.05 and 0.92 ± 0.05 respectively. For PM models, validation accuracies of PM-30, PM-60 and PM-90 reached to 0.92 ± 0.06, 0.90 ± 0.06 and 0.92 ± 0.06 respectively.

**Figure 3:**
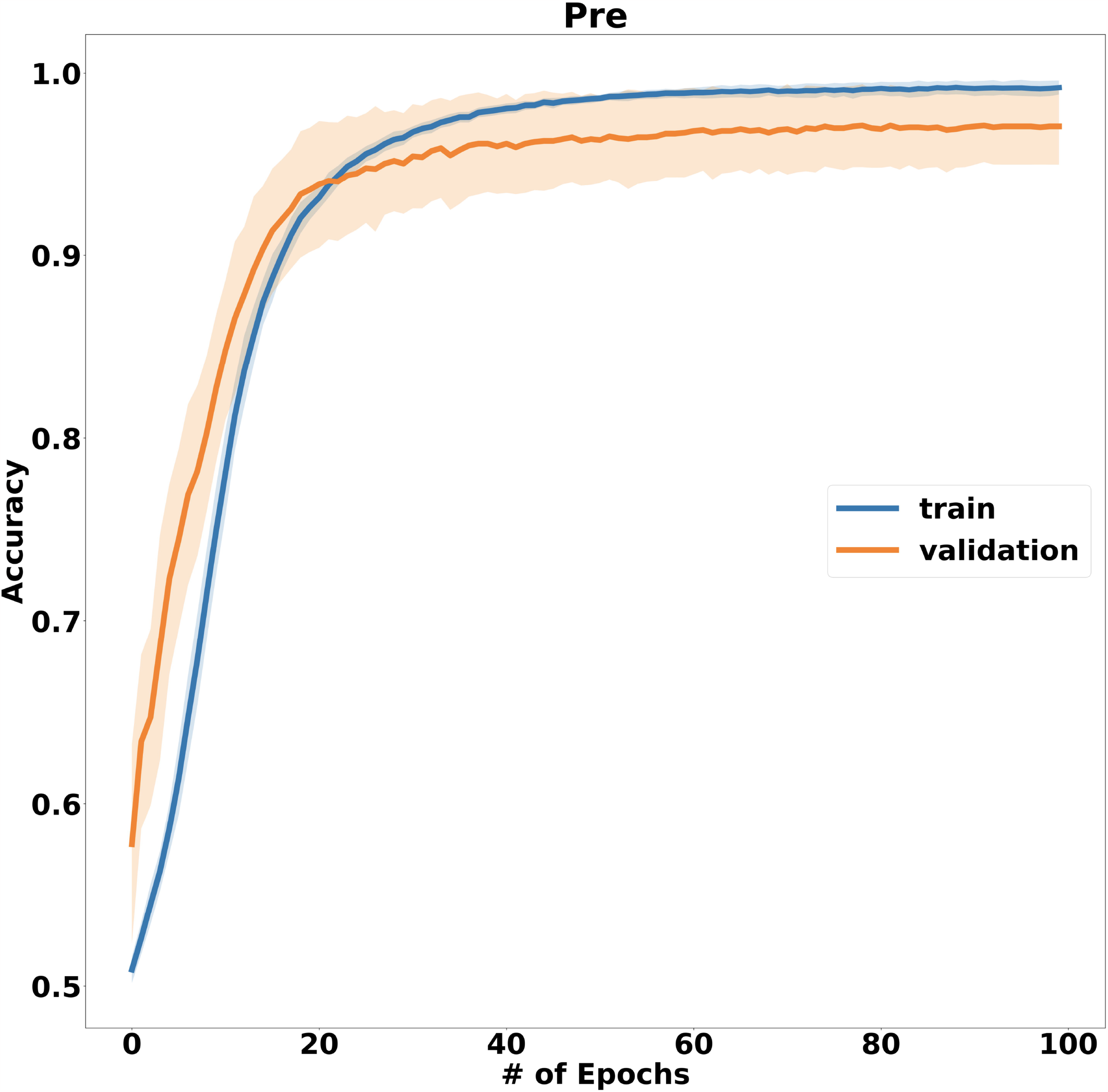
Training and validation accuracy of PDM.

**Figure 4:**
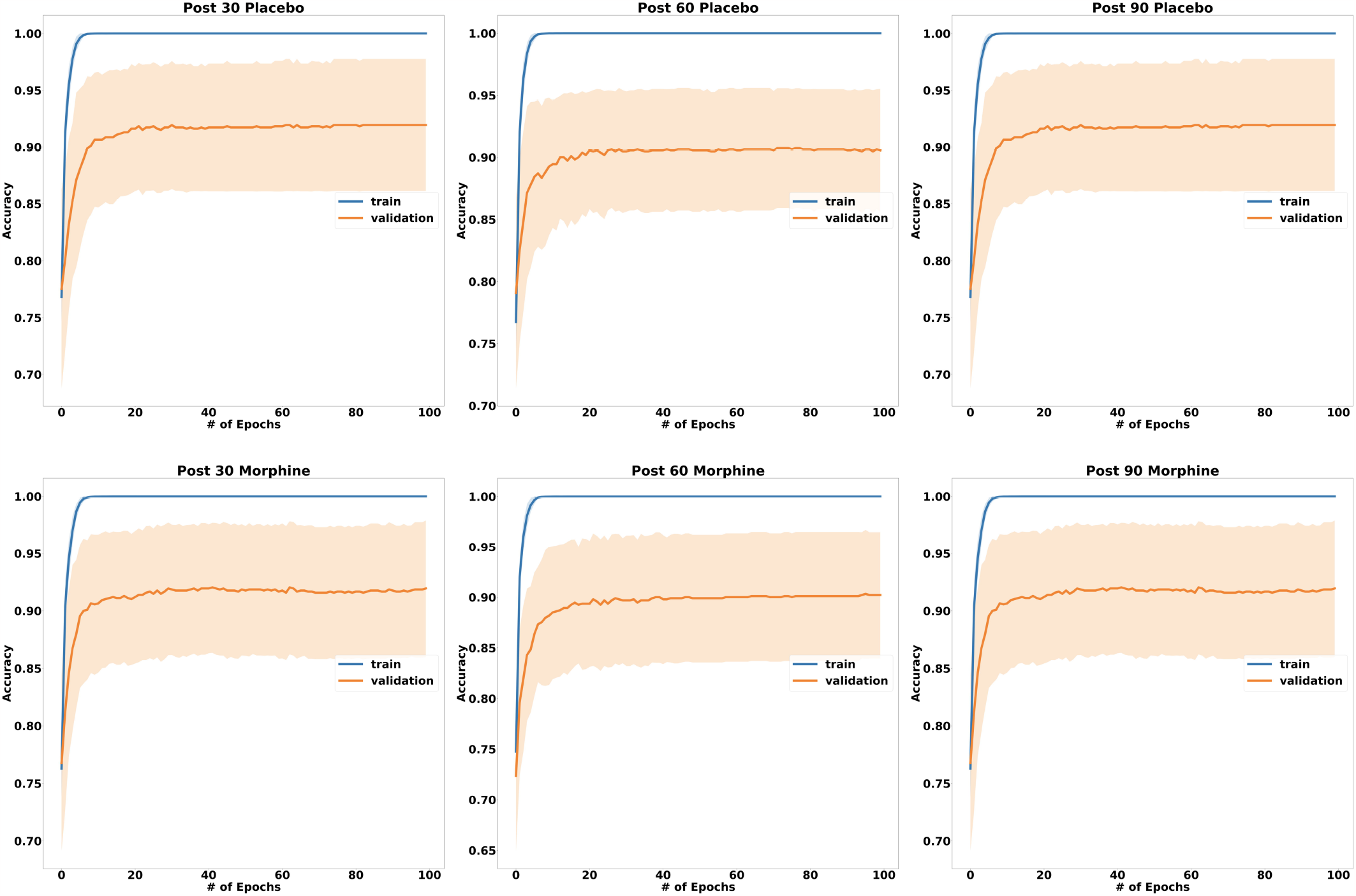
Training and validation accuracy curves of post drug models.

Test performances of PDM and all post-drug models in terms of their accuracy, sensitivity and specificity results are given in Table 2. For PDM, decoding accuracy, sensitivity and specificity performances reached 0.97 ± 0.03, 0.97 ± 0.04, 0.97 ± 0.04 respectively.

Among post-drug morphine models, PM-30 model achieved accuracy, sensitivity and specificity as 0.91 0.05, 0.90 0.08, 0.91 0.07 respectively. For PM-60 model, accuracy, sensitivity and specificity were found as 0.90 0.07, 0.88 0.11, 0.91 0.11 respectively. For PM-90 model, accuracy, sensitivity and specificity were found as 0.91 0.05, 0.89 0.08, 0.92 0.08. respectively. Among post-drug placebo models, accuracy, sensitivity and specificity of PP-30 model were found as 0.92 ± 0.06, 0.92 ± 0.08, 0.91 ± 0.08 respectively. For PP-60 model, accuracy, sensitivity and specificity scores were found as 0.92 ± 0.05, 0.91 ± 0.08, 0.92 ± 0.07 and PP-90 showed accuracy, sensitivity and specificity performance as 0.91 ± 0.07, 0.91 ± 0.08, 0.92 ± 0.10 respectively. Figure 5-7 presents violin plots of accuracy, sensitivity and specificity performances of all models respectively.

**Figure 5:**
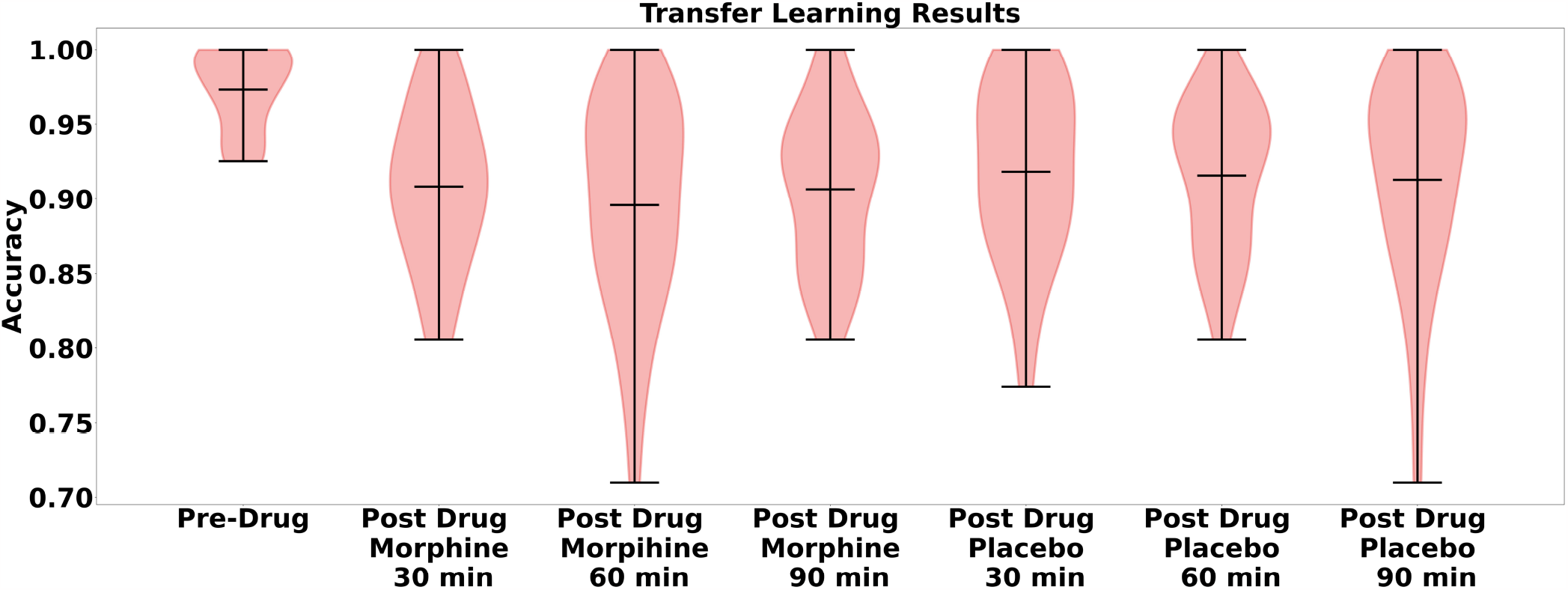
Violin plot of accuracy results of PDM, PP and PM models.

**Figure 6:**
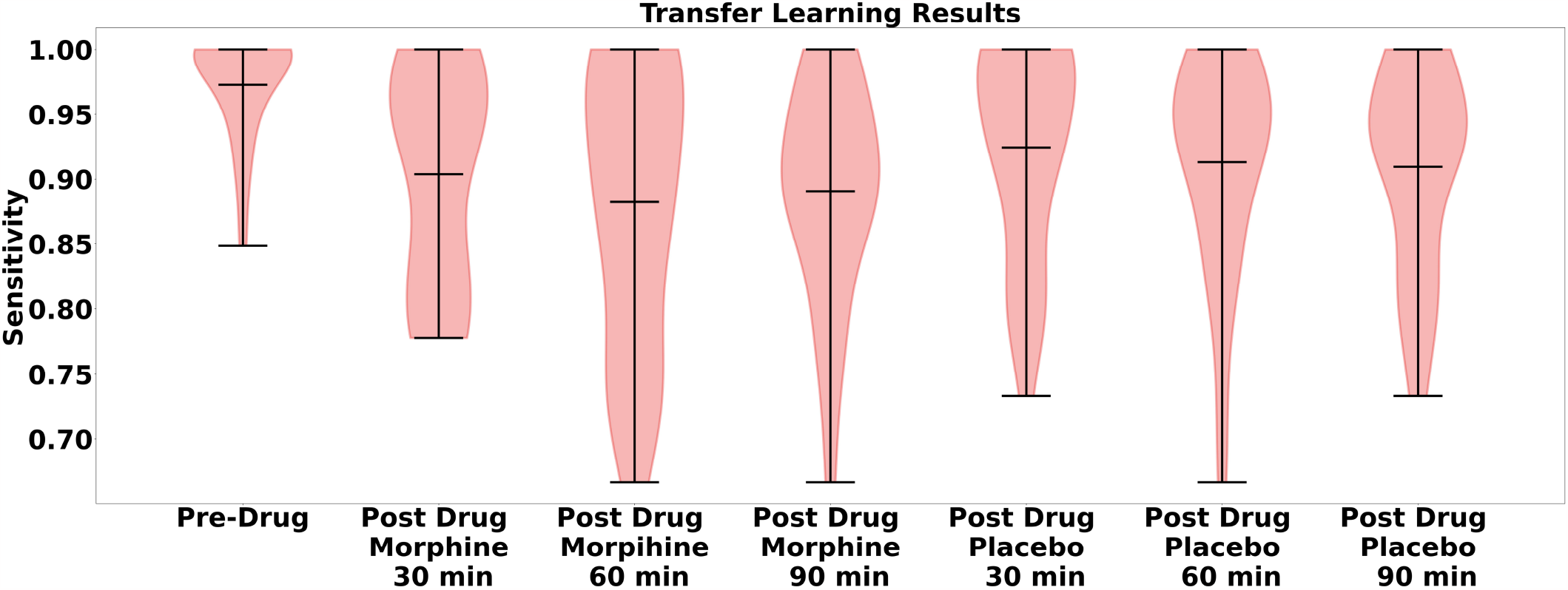
Violin plot of sensitivity results of PDM, PP and PM models.

**Figure 7:**
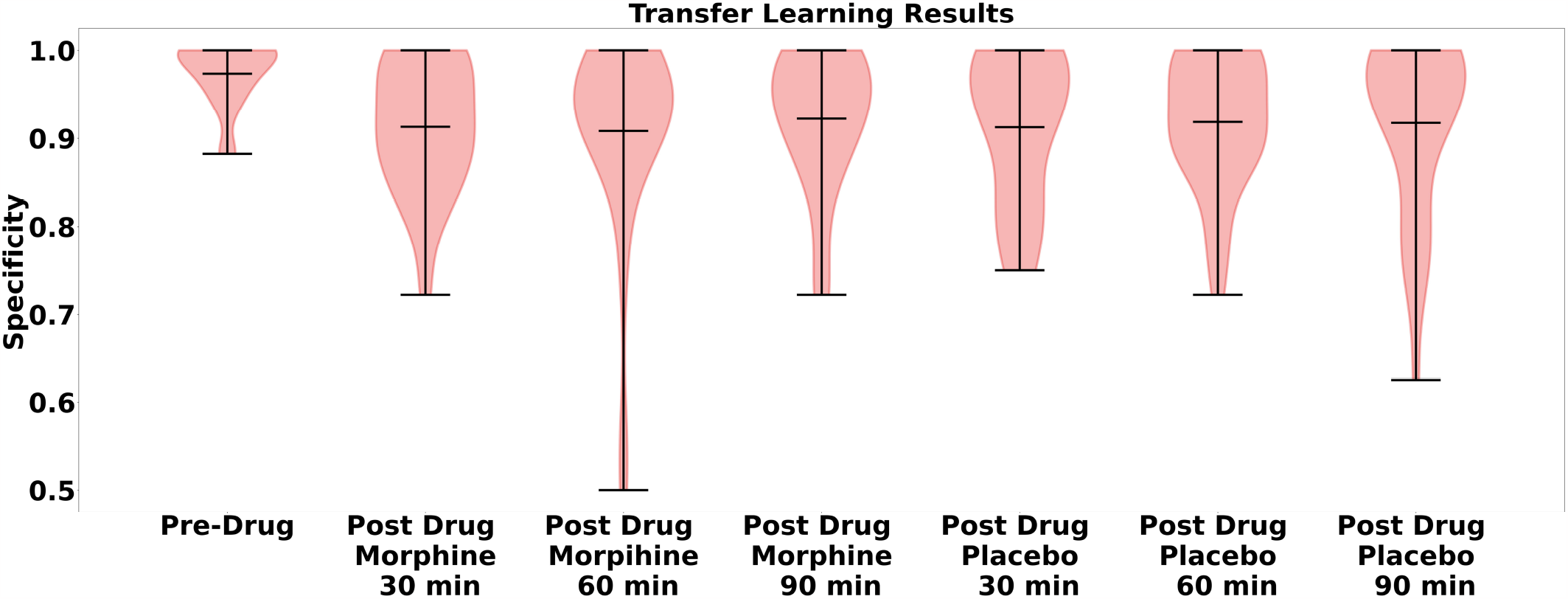
Violin plot of specificity results of all PDM, PP and PM models.

ROC curves of all models are shown in Figure 8. For PDM, AUC was found as 0.97 ± 0.03. For PM models, AUC of PM-30, PM-60 and PM-90 were found as 0.91 ± 0.05, 0.89 ± 0.08 and 0.90 ± 0.06. For PP models, AUC of PP-30, PP-60 and PP-90 were found as 0.92 ± 0.05, 0.92 ± 0.05 and 0.92 ± 0.06.

**Figure 8:**
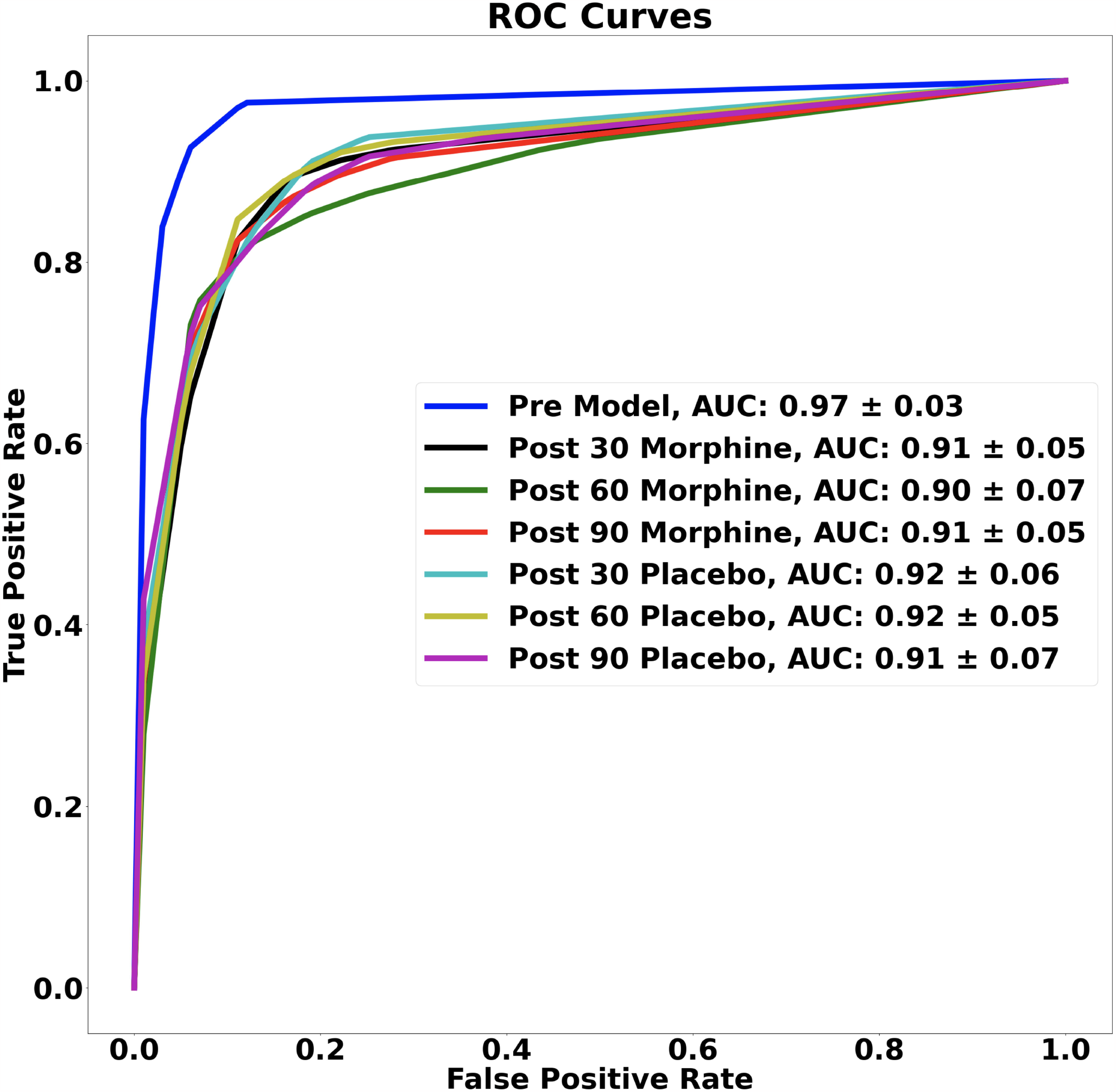
ROC curves and corresponding AUC values of all models.

### 3.2. Statistical Comparison of Model Performances

#### 3.2.1. Pre Drug Model vs Post Drug Models

Kruskal-Wallis test results showed that there is a significant difference between accuracy scores of PDM and post morphine models (χ2(3) = 35.41, p < 0.001). Multiple comparison test using Bonferroni correction showed that PDM accuracy results were significantly higher than PM-30 (mean= 41.55, p<0.001), PM-60 (mean= 45.66, p<0.001) and PM-90 (mean=42.91, p=0.000). No statisticaly significant difference in accuracy performance were found across post-morphine models. Comparison of sensitivity scores between models revealed that there is a significant difference between PDM and PM (χ2(3) = 20.04, p < 0.001). Multiple comparison test using Bonferroni correction showed that PDM sensitivity results were significantly higher than PM-30 (mean= 27.30, p=0.010), PM-60 (mean= 34.05, p<0.001) and PM-90 (mean=33.45, p=0.000). Comparison of specificity scores between PDM and PM revealed that there is a significant difference between models (χ2(3) = 15.68, p<0.001). Multiple comparison test using Bonferroni correction showed that PDM specificity results were significantly higher than PM-30 (mean= 27.30, p=0.006) and PM-60 (mean= 34.05, p=0.002) models but not PM-90 (mean=21.83, p=0.06). No significant difference were found across post-morphine models for any of the performance metrics.

Performance comparison between PDM and post-placebo models revealed that there is a statistically significant difference between accuracy results (χ2(3) = 27.24, p < 0.001). Multiple comparison test using Bonferroni correction showed that PDM accuracy results were significantly higher than PP-30 (mean= 37.06, p=0.00), PP-60 (mean= 38.07, p=0.00) and PP-90 (mean= 38.05, p=0.00). Comparison of sensitivity scores between PDM and post placebo models revealed that there is a significant difference between models (χ2(3) = 15.13, p = 0.001). PDM sensitivity results were significantly higher than post-placebo models (PDM vs PP-30 (mean= 24.23, p=0.028), PDM vs PP-60 (mean= 24.53, p=0.026), PDM vs PP-90 (mean=31.90, p=0.001). Comparison of specificity scores between models revealed that there is a significant difference between PDM and post-placebo models (χ2(3) = 11.44, p = 0.009). PDM specificity results were significantly higher than PP-30 (mean=26.10, p=0.014) and PP-60 (mean= 22.93, p=0.041) models but not PP-90 (mean=22.30, p=0.051). No significant difference were found across post-placebo models for any of the performance metrics.

#### 3.2.2. Classification Performance Comparison of Post Drug Models

2 x 3 repeated measures ANOVA for accuracy scores revealed a significant main effect of drug type (F(1,179)=9.98, p=0.002). Statistical significance for neither the main effect of time condition (F(2,179)=0.1, p=0.901) nor the interaction between drug and time conditions (F(2,179)=0.65, p=0.525) were found. Multiple comparisons for the main effect of drug type revealed that placebo condition showed significantly higher accuracy performance than morphine condition (mean diff. : -0.0198, p = 0.002). 2 x 3 repeated measures ANOVA for sensitivity scores revealed no significance for the main effect of drug type (F(1,179)=3.22, p=0.070), time (F(1,179)=0.62, p=0.540), or a drug and time interaction (F(1,179)=0.08, p=0.922). For specificity results, similarly no significant main effect for drug type (F(1,179)=0.03, p=0.859), time (F(1,179)=0.1, p=0.901) or a drug and time interaction (F(1,179)=0.23, p=0.791) were found.

#### 3.2.3. Shapley Interpretation

Average Shapley values contributed to pre-drug and all post-drug models are shown for every region in Figure 9 and Shapley maps are shown in Figure 10. Positive and negative Shapley values of a region are indicative of positively or negatively contribution of that region to the general decoding performance of the neural processing of two pain levels. Our results showed that, for pre-model R PMC, R DLPFC, L FPA and R MI contributed to decoding pain before drug administration. For PP models, regions that positively contributed to decode pain were; for PP-30 model, L PMC, R PMC, L DLPFC, R FPA and L IFG, for PP-60 model, L PMC, R PMC, R DLPFC, L DLPFC, L FPA R FPA for PP-90 model, L PMC, L DLPFC, L FPA, L IFG, R SMG and R SI.

**Figure 9:**
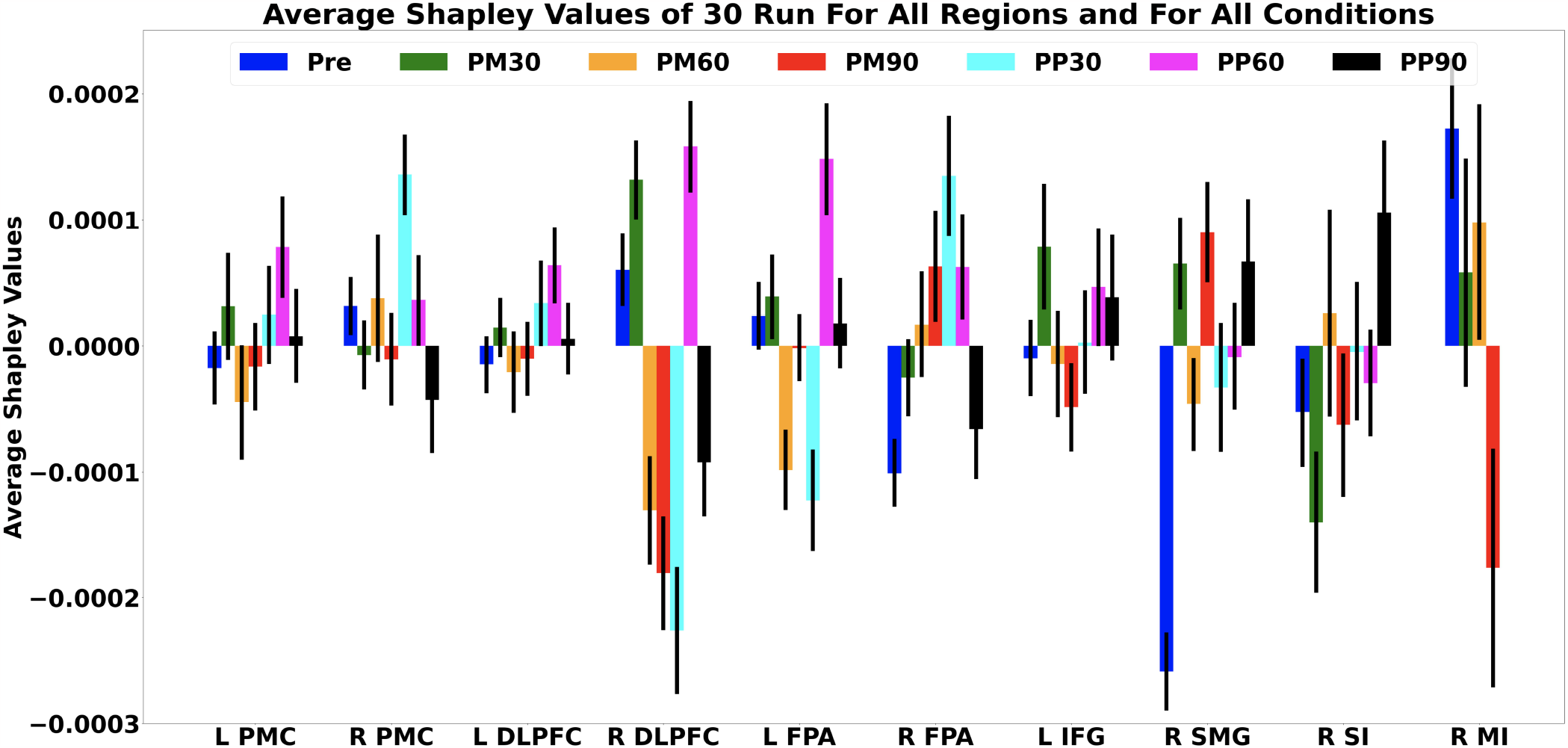
Bar plot of average Shapley values of regional contributions to all models. Pre: Pre-model, PM30 : Post-morphine 30 min, PM60: Post-morphine 60 min, PM90 : Post-morphine 90 min, PP30: Post-Placebo 30 min, PP60: Post-Placebo 60 min, PP90: Post-Placebo 90 min. L: Left, R: Right, PMC: Pre-motor cortex, DLPFC: Dorsolateral Pre Frontal Cortex, FPA: Frontopolar area, IFG: Inferior Frontal Gyrus. SMG: Supramarginal Gyrus, SI: Somatosensory cortex, MI: Motor Cortex

**Figure 10:**
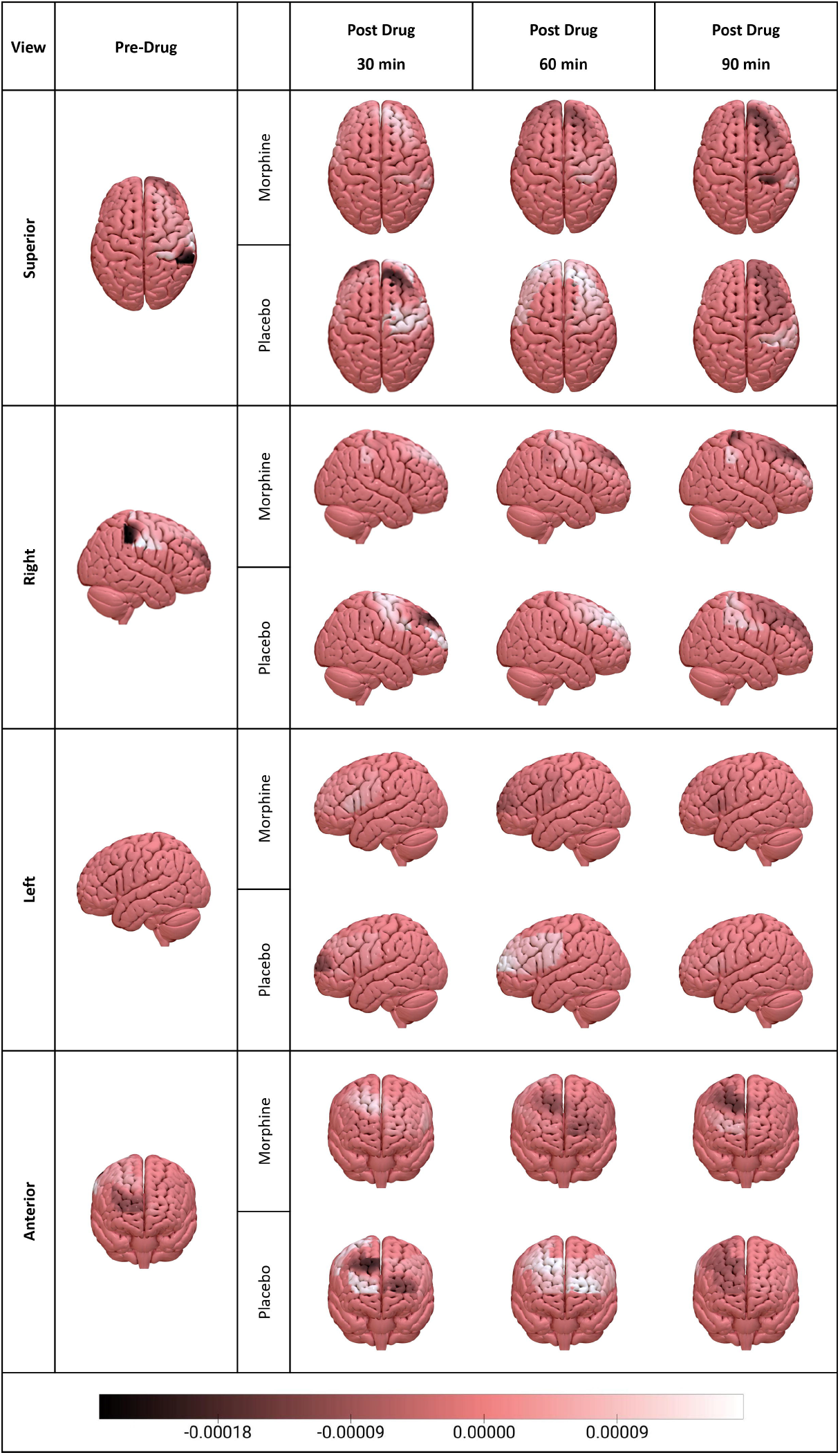
Shapley contributions of regions to corresponding models over the cortex from three axes.

For PM models, regions that positively contributed to decode pain were; for PM-30 model, L PMC, L DLPFC, R DLPFC, L FPA, L IFG, R SMG, and R MI, for PM-60 model, R PMC, R FPA, R SI and R MI and for PM-90 model, R FPA and R SMG.

## 4. Discussion

### 4.1. General Comments and Novelty Emphasis

The aim of this study was to propose a TL based DL methodology for accurate detection and objective classification of the neural processing of painful and non-painful stimuli that were presented under different levels of analgesia. As a relatively new approach for developing neural decoding models (Peterson et al., 2021) for BCI applications (Azab et al., 2018), TL is based on the premise that knowledge generated from a pre-trained base model can be utilized to solve another similar classification problem on a novel data set. Within the context of proposed work, the TL approach was utilized to transfer knowledge (i.e., weights) of the constructed DL model from pre-drug fNIRS scans and the base neural network knowledge of the pre-drug DL model was adapted to the problem of two class classification of the neural processing of two levels of nociceptive stimuli (Peng, Yucel, et al., 2018). The motivation behind utilizing a TL approach relied on the assumption that such an adaptive training methodology would demonstrate a high performance while being computationally efficient and would remove the necessity to build new DL models for data collected at different clinical or daily life conditions for which obtaining training data is not practical and building a new model will have a computational cost. The feasibility of TL approach was demonstrated by its efficacy in predicting the class of two levels of nociceptive stimuli from noninvasive fNIRS recordings obtained under two different pharmacological conditions and at three time points post-intervention. Each of the post-drug models had mean accuracy, sensitivity, specificity and AUC performance above 90% when the weights obtained from the base model were transferred and no statistically significant difference in classification performance were found across the post-drug models for any of the performance metrics. These results demonstrated that knowledge obtained from a pre-drug base model could be successfully utilized to build novel models for predicting the perceived pain intensity level from neurally induced hemodynamic signals obtained at 6 distinct dynamic brain states that were altered with either analgesic or a placebo intervention and at 3 different times post-drug administration.

The presented work includes several novelties. First, to the best of our knowledge, this is the first fNIRS based pain decoding approach in healthy subjects before and after medication. To date, there have been no studies that have demonstrated the efficacy of TL methodology for single trial classification of the presence of painful stimuli processing under different pharmacological conditions that included analgesics. Second, this is the first study that have tested the feasibility of integrating TL methodology with neurophysiological data obtained from a noninvasive, mobile and wearable fNIRS system for the purpose of predicting the perceived pain intensity level under different drug administrations in healthy male adults. This approach was quite remarkable because it provides a proof of concept preliminary analysis that demonstrates the practicality of adapting a pre-drug base decoding model to different clinical conditions where collecting training data is not possible. This is the first study that explains the behavior of a pain decoding model both at pre-drug and post-drug conditions by utilizing an explainable AI approach where the motivation was to understand which cortical regions contributed to the output of model at most for every session.

### 4.2. Comparison of the Classification Performances of PDM and Post Drug Models

Test performances of PDM and all post-drug models achieved mean accuracy, specificity, sensitivity and AUC scores above 90%. Mean performance of PDM was above 95% for accuracy, sensitivity, specificity and AUC metrics. PDM had a statistically significantly higher 2 class classification performance than all PM models for all performance metrics while PM models did not demonstrate a significant difference among each other for any of the metrics. Similarly, PDM had a statistically significantly higher 2 class classification performance than all PP models for all metrics while PP models did not demonstrate a significant difference among each other for any of the metrics.

Although the base model has a higher performance that post drug models, the fact that all models have a general classification performance above 90 % in all performance metrics demonstrate that knowledge obtained from a pre-drug base model could be successfully utilized to build novel models for predicting the pain intensity level from neurally induced hemodynamic signals. Performance of PM models were relatively lower than that of PDM and PP models. This result is expected as morphine alters hemodynamic response patterns in several cortical regions including MPFC as shown in the previous work of Peng et al (2018) from whom the dataset was obtained.

Neurophysiological and behavioral consequences of pharmacological interventions during acute (Barkin & Barkin, 2001; Eland, 1988; Jeha et al., 2021) and chronic pain (Coles et al., 2022; Kuijpers et al., 2011; Park & Moon, 2010) conditions have been thoroughly examined in the last few decades. Recently, neural correlates of various drug interventions have been investigated by using fMRI (Hansen et al., 2015; Tinnermann et al., 2022; Wager et al., 2013) and fNIRS (Peng et al., 2021; Peng, Yucel, et al., 2018). Peng and colleagues conducted an fNIRS study that focused on the effect of placebo and morphine intervention on neural correlates of acute pain and they reported that morphine attenuates hemodynamic response activity in the medial frontopolar area (Peng, Yucel, et al., 2018). In addition, Hansen and colleagues conducted an fMRI study that examined the neural effects of analgesic drugs (morphine / placebo) under acute painful stimulation and while morphine-based attenuation was observed in the right insula, anterior cingulate cortex and inferior parietal cortex, no difference in brain activation between pre and post placebo administration conditions was observed (Hansen et al., 2015). These studies highlighted the fact that morphine administration alters cortical hemodynamic activity induced by neural processing of nociceptive stimuli. One potential reason for the relatively lower classification performance of PM models may be due to the alterations of hemodynamic responses obtained during both pain and non-pain conditions in several brain regions with morphine administration with respect to the neural activity obtained during drug-free condition. Wager and colleagues conducted a study that focused on extracting a pain signature. They combined fMRI measures with a machine learning method (least absolute shrinkage and selection operator regularized principal components regression – LASSO PCR) to classify the neural processing of painful and non-painful stimuli under remifentanil administration. Their proposed methodology achieved 90% sensitivity and 81 % specificity before drug administration and 86 % sensitivity and 62% specificity were achieved after drug treatment (Wager et al., 2013). The study whose dataset was utilized in the presented work also found that morphine reduced the pain induced hemodynamic responses in MPFC however; it did not change the responses induced by non-painful stimuli. Meanwhile, placebo drug affected the spatiotemporal patterns of neither painful nor non-painful induced hemodynamic responses (Peng, Yucel, et al., 2018).

On the other hand, 2 way ANOVA results demonstrated that the accuracy of PP models were significantly greater than the decoding accuracy of PM models. Chen (2021) provides an excellent review on the performance of different supervised and unsupervised classification algorithms in correct identification of acute and chronic pain conditions by use of data obtained from different functional imaging modalities. In our work, the accuracy, sensitivity, specificity performances of post drug models were not statistically significantly different from each other per intervention type while they remained in the high-performance spectrum among the performance metrics reported in previous studies which targeted two class classification of pain intensity by use of functional neuroimaging measures (Chen, 2021).

### 4.3 Interpretation of Regional Shapley Contributions

#### 4.3.1 Pre-Drug Condition

In the pre-drug stage, R PMC, R DLPFC, L FPA and R MI positively contributed to the highly accurate (%97) decoding performance of PDM. Among these regions, MI is a widely-known and key region in pain processing which has a notable role on integrating sensory and motor aspects of pain (Brown et al., 2011; Leknes & Tracey, 2008; Martucci & Mackey, 2018). DLPFC is involved in several cognitive processes such as attention (Bidet-Caulet et al., 2015; Vossel et al., 2014; Voytek et al., 2010) and working memory (Barbey et al., 2013) as well as neural processing of acute and chronic pain (Seminowicz & Moayedi, 2017). Previous acute pain studies revealed that bilateral DLPFC activity has a negative correlation with the extent of unpleasantness of thermal pain (Lorenz et al., 2003) and pain catastrophizing scores (Seminowicz & Davis, 2006). On the other hand, R DLPFC was found to be strongly associated with control of perceived pain intensity (Wiech et al., 2006). The positive contribution of DLPFC to PDM classification performance may be due to its involvement in the above-mentioned cognitive aspects of pain experience.

R PMC (BA 6) was also found to be a positive contributor to the classification performance of PDM. BA6 is a large cortical area located at the anterior side of MI and this region is primarily responsible for motor acts such as writing and speech besides sensory guidance of movement (Tanji, 1996; Wise, 1985). A previous arterial spin labeling-MRI (ASL-MRI) study revealed that acute cold and heat pain resulted in an increase in cerebral blood flow (CBF) (Frolich et al., 2012) which could serve as a potential biomarker of acute pain. In a PET based CBF study, PMC showed significant responses to both heat and cold pain in both genders (Casey, 1999). However, how PMC is effective in pain processing still remains unclear. Previous studies claimed that activity increase in PMC might be related to anticipation of movements to avoid painful stimuli (Hsieh et al., 1994). Contribution of R PMC to classification performance of our PDM model might be related to its role in regulating avoidance behavior for painful and non-painful stimuli.

L FPA (BA 10), a region located at the anterior portion of the PFC, was previously found to be a critical region in pain processing in previous fMRI (Cauda et al., 2010; Hautvast et al., 1997; Kucyi et al., 2014; Lobanov et al., 2013; Porro et al., 1998; Svensson et al., 1997) and fNIRS (Aasted et al., 2016; Peng, Yucel, et al., 2018) studies. Previous reports suggest that FPA might be involved in collation, integration and high-level processing of pain (Peng, Steele, et al., 2018). In the study from which the dataset was generated, statistically significant difference in hemodynamic responses to painful and non-painful stimuli was found in medial BA 10 of the pre-scan datasets of morphine and placebo visits of all subjects (Peng, Yucel, et al., 2018). The significant difference in hemodynamic responses to painful and non-painful stimuli might be the reason for positive contribution of L FPA to the highly accurate PDM decoding performance. The role of BA 10 in pain perception still remains unclear. However, anatomical connections exist between BA 10 and several cortical and subcortical regions such as thalamus, insula (Burman et al., 2011; Petrides & Pandya, 2007) and anterior cingulate cortex (ACC) (Bushnell et al., 2013; Coghill et al., 2003; Derbyshire et al., 1994) which play important roles in sensory-discrimination and pain perception (Peng, Steele, et al., 2018).

#### 4.3.2 Post-Drug Condition

Among PP models, L PMC, R PMC, L DLPFC, R FPA and L IFG positively contributed to the classification performance of PP-30. L PMC, R PMC, R DLPFC, L DLPFC, L FPA, R FPA regions positively contributed to the classification performance of PP-60 and L PMC, L DLPFC, L FPA, L IFG, R SMG and R SI positively contributed to the classification performance of PP-90 model. Regions that contributed both to PDM and PP models were R PMC and R DLPFC for PP-30, R PMC and L FPA for PP-60 and L FPA for PP-90.

Common positively contributing regions to the classification performance of PDM and PM models were R PMC for PM-30, R PMC, R DLPFC, L FPA for PM-60 and L FPA for PM-90. Despite these positive contributor regions common to both PDM and PM models, additional regions also contributed to the output of the PM models. This observation may suggest that transferring knowledge from a pre-drug base model might be useful to decode the presence of a painful response, however information from additional cortical regions may also be needed for a high decoding performance in post-drug models because of the intra and intersubject variability introduced to fNIRS signals by efficacy duration of analgesic drug.

A recent fMRI meta-analysis on placebo analgesia revealed that placebo administration causes small and widespread activity reductions during painful stimuli processing in several brain regions that are related to both painful stimulus and decision-making processes (Zunhammer et al., 2021). DLPFC and PMC were found to be the common contributor regions across all developed models for PP models. Among these regions, PMC and SMA were previously reported as critical regions which might reflect placebo effect on pain-induced hemodynamic response (Frolich et al., 2012). PMC activation was reported during painful stimulation under high level of placebo administration (J. C. Choi et al., 2011). In the same study, a positive correlation was found between PMC and ACC activities and ACC activity is strongly related to placebo and opioid analgesia (Bingel et al., 2006; Petrovic et al., 2002). Changes in the hemodynamic activity in PMC might be associated with the hemodynamic activity in ACC which cannot be measured by using fNIRS. On the other hand, DLPFC plays a role in pain suppression by attention-based pain regulation (Graff-Guerrero et al., 2005; Krummenacher et al., 2010; Lorenz et al., 2003; Peyron et al., 2000) and it was particularly involved in placebo analgesia (Pariente et al., 2005; Wager et al., 2004). Previous studies also reported that there was a correlation between DLPFC connectivity and placebo analgesia (Tetreault et al., 2016; Vachon-Presseau et al., 2018). We think that DLPFC positively contributed to pain decoding during placebo analgesia due to its pain regulatory role.

Similarly, FPA (BA 10) was also found to be another region that positively contributed to decoding performance of PP models. Previous evidence suggests that FPA plays a role in pain anticipation under placebo analgesia (Amanzio et al., 2013; Petrovic et al., 2010; Watson et al., 2009) and increased activation in FPA during pain expectation might be related to placebo analgesia and emotional regulation (Amanzio et al., 2013). Compared to Amanzio et al (2013), L IFG presented a placebo-induced activation increase in another meta-analysis (Atlas & Wager, 2014) and is considered as a critical anticipatory predictor of placebo analgesia (Wager et al., 2011). We think that both these regions contributed to decoding performance due to their regulation and anticipation roles during placebo analgesia condition. Previous evidence related to behavior of SI showed that pain-induced activity decreased after placebo analgesia (Bingel et al., 2006; Eippert et al., 2009; Lui et al., 2010). However, in the previous study of this dataset, no significant difference was reported in SI between painful and non-painful stimuli when compared to pre-drug status (Peng, Yucel, et al., 2018). In that study, due to not having any comparison between painful and non-painful stimuli for each drug condition, it is hard to make a direct interpretation related to the reason of contribution of SI. Statistical similarity does not fully guarantee a low accurate discrimination of two classes by using ML approaches (Arbabshirani et al., 2017). On the other hand, SMG is located at the inferior parietal lobule (IPL) which is involved in pain relief (Jae Chan Choi et al., 2022; Zunhammer et al., 2021). Wager and colleagues found that SMG is a positive predictor of decoding painful vs. non-painful stimulus (Wager et al., 2013).

For PM models, regions that positively contributed to decoding performance were; L PMC, L DLPFC, R DLPFC, L FPA, L IFG, R SMG and R MI positively contributed to the model after 30 min of administration. R PMC, R FPA, R SI and R MI positively contributed to the model after 60 min of administration and R FPA and R SMG positively contributed to the model after 90 min of administration. Effects of opioids like morphine and its derivatives such as remifentanil on painful stimulus have previously been investigated in several fMRI (Becerra et al., 2006; Gear et al., 2013; Hansen et al., 2015; Wager et al., 2013; Wanigasekera et al., 2012) and fNIRS (Peng, Yucel, et al., 2018) studies. Morphine induced activation reduction was observed in DLPFC (Becerra et al., 2006), inferior parietal lobe which covers SMG (Becerra et al., 2006; Hansen et al., 2015) and FPA (Peng, Yucel, et al., 2018). Among these regions, a previous MR-spectroscopy study revealed that frontal region is an opioid rich region (Hansen et al., 2016) which is possibly effective in reducing perceived pain intensity. On the other hand, while pain-induced hemodynamic activity reduction in SI was observed after morphine administration (Gear et al., 2013; Peng, Yucel, et al., 2018), no difference was found between pre-morphine and post-morphine non-painful stimuli induced hemodynamic activity in SI (Becerra et al., 2006; Peng, Yucel, et al., 2018).

### 4.3. Potential of the Proposed Methodologies

The presented work demonstrated that knowledge obtained from a pre-drug base model could be successfully transferred to build novel models for predicting the perceived pain intensity level from neurally induced hemodynamic signals obtained at 6 distinct dynamic brain states which were altered with either analgesic or a placebo intervention and at 3 different times post-drug administration. We provide a proof of concept preliminary analysis that demonstrates the practicality of adapting a pre-drug base decoding model to different clinical conditions where collecting training data is not possible. The low computational cost and high classification performance of TL approach makes it feasible for specific classification problems where a baseline data is available and a model trained with this baseline data can be adapted to data collected at different clinical or daily life conditions where obtaining training data is not feasible/practical to build novel ML or DL models.

Unveiling the explanation power of features obtained from different cortical regions of interest is prominent as it may aid the design of more computationally efficient BCI system designs that target pain detection and such an approach may provide more precisely localized physiological markers of pain. In the presented work, Shapley values presented no consistent localization of positive contribution across all models. Nonetheless, the proposed combination of TL based DL methodology with an xAI method and their application to fNIRS data demonstrate a potential for unveiling the explanation power of different ROIs and this analytical procedure may aid the design of more computationally efficient BCI system designs for other application areas.

### 4.4. Limitations of the Study and Recommendations for Future Work

Pain is a multisensory experience and test retest reliability is always questionable in human functional neuroimaging studies that target cognitive and emotional aspects. We should not ignore the fact that pain responsive cortical areas do not solely process pain induced neural information. Both morphine and placebo interventions result in different cognitive and anticipation effects and decoding the intensity of a painful stimulus and its saliency dimension cannot be differentially performed (Lee et al. 2020). Due to these constraints, we should take into account the fact that the relative contribution of morphine and placebo modulated regions may show variability within and across participants and across different pharmacological conditions. Painful and non-painful stimuli may have different hemodynamic activation strengths at each post drug session which may be not only due to the differential effect of the type of drug administration but also due to the varying cognitive state at each session including habituation effects.

Although the number of subjects in our study was comparable to the sample sizes reported in previous pain decoding studies, low number of subjects was another critical limitation in our study. DL algorithms require high amounts of data for training (Szucs & Ioannidis, 2020). However, obtaining comparably high numbers of labeled data in medicine field is difficult due to factors such as acquisition cost and labor. Hence, clinical studies are conducted with relatively limited amount of data when compared to other areas of DL applications. To overcome this limitation, we applied a data augmentation procedure during model training by utilizing well accepted data augmentation approaches (Wen et al., 2021). Nonetheless, while synthetic data are expected to capture the diversity and variability available in real-world data, its creation is still a biased approach. For best case scenarios, training DL algorithms with more real world data from more participants will increase the reliability of validation and reproducibility of our results. Besides, although DL methodologies remove the necessity of feature engineering and domain knowledge requirements, it should not be neglected that they still have many unknown parameters and require vast amounts of labelled samples for training.

## 5. Conclusion

The presented work addressed two main research questions. Our first question aimed to assess the feasibility of implementing a TL methodology to decode the neural processing of of painful and non-painful stimuli obtained under two distinct pharmacological interventions and at different post-intervention times. Our results demonstrated that the neural processing of painful and non-painful stimuli states could be successfully distinguished by utilizing hemodynamic information obtained before and after a morphine or a placebo drug administration. The performance of the TL approach in accurate classification of pain intensity level was tested on 6 distinct post models which were fine-tuned for fNIRS data recorded during noxious and innocious stimuli under different pharmacological conditions. Our results demonstrated the potential of training models with a baseline fNIRS data and adapting these baseline models to data collected at different clinical or daily life conditions where obtaining training data is not feasible/practical to build novel ML or DL models. Our second aim was to assess the contribution of features obtained from different cortical regions to the classification performance of the proposed DL model and how this contribution changes as hemodynamic activity is modified with morphine or placebo intervention. Our findings demonstrate the potential of proposed methodology for unveiling the explanation power of different ROIs and how this approach may aid the design of more computationally efficient fNIRS based BCI system designs for other daily-life and clinical application areas.

## Supporting information

Highlights

Data and Code Availability

Declaration of Interest

## Data Availability

In addition to HomER3, Tensorflow toolkit (version 2.8.0) and Shap toolbox, the code can be downloaded from the website https://github.com/aykuteken/Pain_decoding/ The data information can be found in https://www.nitrc.org/projects/yucel18pain/.

## CRediT authorship contribution statement

Aykut Eken: Conceptualization, Investigation, Methodology, Formal Analysis, Software, Validation, Visualization, Writing – original draft, review & editing. Sinem Burcu Erdoğan: Investigation, Writing – original draft, review & editing. Gülnaz Yükselen: Investigation, Visualization, Murat Yüce: Investigation, Validation. All authors have read and agreed to the submitted version of the manuscript.

## References

Aasted, C. M., Yucel, M. A., Steele, S. C., Peng, K., Boas, D. A., Becerra, L., & Borsook, D. (2016). Frontal Lobe Hemodynamic Responses to Painful Stimulation: A Potential Brain Marker of Nociception. PLoS One, 11(11), e0165226. doi:10.1371/journal.pone.0165226

Abadi, M. n., Barham, P., Chen, J., Chen, Z., Davis, A., Dean, J., … Zheng, X. (2016). TensorFlow: A System for Large-Scale Machine Learning. Osdi’16, 265–283.

Amanzio, M., Benedetti, F., Porro, C. A., Palermo, S., & Cauda, F. (2013). Activation likelihood estimation meta-analysis of brain correlates of placebo analgesia in human experimental pain. Hum Brain Mapp, 34(3), 738–752. doi:10.1002/hbm.21471

Arbabshirani, M. R., Plis, S., Sui, J., & Calhoun, V. D. (2017). Single subject prediction of brain disorders in neuroimaging: Promises and pitfalls. Neuroimage, 145(Pt B), 137–165. doi:10.1016/j.neuroimage.2016.02.079

Atlas, L. Y., & Wager, T. D. (2014). A meta-analysis of brain mechanisms of placebo analgesia: consistent findings and unanswered questions. Handb Exp Pharmacol, 225, 37–69. doi:10.1007/978-3-662-44519-8_3

Azab, A. M., Toth, J., Mihaylova, L. S., & Arvaneh, M. (2018). A review on transfer learning approaches in brain–computer interfaceSignal Processing and Machine Learning for Brain-Machine InterfacesControl, Robotics & Sensors (pp. 81–101): Institution of Engineering and Technology. Retrieved from https://digital-library.theiet.org/content/books/10.1049/pbce114e_ch5. doi:10.1049/PBCE114E_ch5

Barbey, A. K., Koenigs, M., & Grafman, J. (2013). Dorsolateral prefrontal contributions to human working memory. Cortex, 49(5), 1195–1205. doi:10.1016/j.cortex.2012.05.022

Barkin, R. L., & Barkin, D. (2001). Pharmacologic management of acute and chronic pain: focus on drug interactions and patient-specific pharmacotherapeutic selection. South Med J, 94(8), 756–770.

Becerra, L., Aasted, C. M., Boas, D. A., George, E., Yucel, M. A., Kussman, B. D., … Borsook, D. (2016). Brain measures of nociception using near-infrared spectroscopy in patients undergoing routine screening colonoscopy. Pain, 157(4), 840–848. doi:10.1097/j.pain.0000000000000446

Becerra, L., Harter, K., Gonzalez, R. G., & Borsook, D. (2006). Functional magnetic resonance imaging measures of the effects of morphine on central nervous system circuitry in opioid-naive healthy volunteers. Anesth Analg, 103(1), 208-216, table of contents. doi:10.1213/01.ane.0000221457.71536.e0

Bidet-Caulet, A., Buchanan, K. G., Viswanath, H., Black, J., Scabini, D., Bonnet-Brilhault, F., & Knight, R. T. (2015). Impaired Facilitatory Mechanisms of Auditory Attention After Damage of the Lateral Prefrontal Cortex. Cereb Cortex, 25(11), 4126–4134. doi:10.1093/cercor/bhu131

Bingel, U., Lorenz, J., Schoell, E., Weiller, C., & Buchel, C. (2006). Mechanisms of placebo analgesia: rACC recruitment of a subcortical antinociceptive network. Pain, 120(1-2), 8–15. doi:10.1016/j.pain.2005.08.027

Bornhovd, K., Quante, M., Glauche, V., Bromm, B., Weiller, C., & Buchel, C. (2002). Painful stimuli evoke different stimulus-response functions in the amygdala, prefrontal, insula and somatosensory cortex: a single-trial fMRI study. Brain, 125(Pt 6), 1326–1336. doi:10.1093/brain/awf137

Brown, J. E., Chatterjee, N., Younger, J., & Mackey, S. (2011). Towards a physiology-based measure of pain: patterns of human brain activity distinguish painful from non-painful thermal stimulation. PLoS One, 6(9), e24124. doi:10.1371/journal.pone.0024124

Burman, K. J., Reser, D. H., Richardson, K. E., Gaulke, H., Worthy, K. H., & Rosa, M. G. (2011). Subcortical projections to the frontal pole in the marmoset monkey. Eur J Neurosci, 34(2), 303–319. doi:10.1111/j.1460-9568.2011.07744.x

Bushnell, M. C., Ceko, M., & Low, L. A. (2013). Cognitive and emotional control of pain and its disruption in chronic pain. Nat Rev Neurosci, 14(7), 502–511. doi:10.1038/nrn3516

Casey, K. L. (1999). Forebrain mechanisms of nociception and pain: analysis through imaging. Proc Natl Acad Sci U S A, 96(14), 7668–7674. doi:10.1073/pnas.96.14.7668

Cauda, F., D’Agata, F., Sacco, K., Duca, S., Cocito, D., Paolasso, I., … Geminiani, G. (2010). Altered resting state attentional networks in diabetic neuropathic pain. J Neurol Neurosurg Psychiatry, 81(7), 806–811. doi:10.1136/jnnp.2009.188631

Chen, Z. S. (2021). Decoding pain from brain activity. J Neural Eng, 18(5). doi:10.1088/1741-2552/ac28d4

Choi, J. C., Park, H.-J., Park, J. A., Kang, D. R., Choi, Y.-S., Choi, S., … Kim, J. (2022). The increased analgesic efficacy of cold therapy after an unsuccessful analgesic experience is associated with inferior parietal lobule activation. Scientific Reports, 12(1), 14687. doi:10.1038/s41598-022-18181-0

Choi, J. C., Yi, D. J., Han, B. S., Lee, P. H., Kim, J. H., & Kim, B. H. (2011). Placebo effects on analgesia related to testosterone and premotor activation. Neuroreport, 22(9), 419–423. doi:10.1097/WNR.0b013e32834601c9

Coghill, R. C., McHaffie, J. G., & Yen, Y. F. (2003). Neural correlates of interindividual differences in the subjective experience of pain. Proc Natl Acad Sci U S A, 100(14), 8538–8542. doi:10.1073/pnas.1430684100

Coles, S., Dabbs, W., & Wild, S. (2022). Pharmacologic Management of Chronic Pain. Prim Care, 49(3), 387–401. doi:10.1016/j.pop.2022.01.005

Cope, M., Delpy, D. T., Reynolds, E. O., Wray, S., Wyatt, J., & van der Zee, P. (1988). Methods of quantitating cerebral near infrared spectroscopy data. Adv Exp Med Biol, 222, 183–189. doi:10.1007/978-1-4615-9510-6_21

De Felice, M., & Ossipov, M. H. (2016). Cortical and subcortical modulation of pain. Pain Manag, 6(2), 111–120. doi:10.2217/pmt.15.63

Derbyshire, S. W., Jones, A. K., Devani, P., Friston, K. J., Feinmann, C., Harris, M., … Frackowiak, R. S. (1994). Cerebral responses to pain in patients with atypical facial pain measured by positron emission tomography. J Neurol Neurosurg Psychiatry, 57(10), 1166–1172. doi:10.1136/jnnp.57.10.1166

Eippert, F., Bingel, U., Schoell, E. D., Yacubian, J., Klinger, R., Lorenz, J., & Buchel, C. (2009). Activation of the opioidergic descending pain control system underlies placebo analgesia. Neuron, 63(4), 533–543. doi:10.1016/j.neuron.2009.07.014

Eland, J. M. (1988). Pharmacologic management of acute and chronic pediatric pain. Issues Compr Pediatr Nurs, 11(2-3), 93–111. doi:10.3109/01460868809038008

Fekete, T., Rubin, D., Carlson, J. M., & Mujica-Parodi, L. R. (2011). The NIRS Analysis Package: noise reduction and statistical inference. PLoS One, 6(9), e24322. doi:10.1371/journal.pone.0024322

Frolich, M. A., Deshpande, H., Ness, T., & Deutsch, G. (2012). Quantitative changes in regional cerebral blood flow induced by cold, heat and ischemic pain: a continuous arterial spin labeling study. Anesthesiology, 117(4), 857–867. doi:10.1097/ALN.0b013e31826a8a13

Gear, R., Becerra, L., Upadhyay, J., Bishop, J., Wallin, D., Pendse, G., … Borsook, D. (2013). Pain facilitation brain regions activated by nalbuphine are revealed by pharmacological fMRI. PLoS One, 8(1), e50169. doi:10.1371/journal.pone.0050169

Graff-Guerrero, A., Gonzalez-Olvera, J., Fresan, A., Gomez-Martin, D., Mendez-Nunez, J. C., & Pellicer, F. (2005). Repetitive transcranial magnetic stimulation of dorsolateral prefrontal cortex increases tolerance to human experimental pain. Brain Res Cogn Brain Res, 25(1), 153–160. doi:10.1016/j.cogbrainres.2005.05.002

Green, S., Karunakaran, K. D., Labadie, R., Kussman, B., Mizrahi-Arnaud, A., Morad, A. G., … Borsook, D. (2022). fNIRS brain measures of ongoing nociception during surgical incisions under anesthesia. Neurophotonics, 9(1), 015002. doi:10.1117/1.NPh.9.1.015002

Gundel, H., Valet, M., Sorg, C., Huber, D., Zimmer, C., Sprenger, T., & Tolle, T. R. (2008). Altered cerebral response to noxious heat stimulation in patients with somatoform pain disorder. Pain, 137(2), 413–421. doi:10.1016/j.pain.2007.10.003

Han, H. (2022). The Utility of Receiver Operating Characteristic Curve in Educational Assessment: Performance Prediction. Mathematics, 10(9). Retrieved from https://mdpi-res.com/d_attachment/mathematics/mathematics-10-01493/article_deploy/mathematics-10-01493-v2.pdf?version=1651465465 doi:10.3390/math10091493

Hansen, T. M., Olesen, A. E., Graversen, C., Drewes, A. M., & Frokjaer, J. B. (2015). The Effect of Oral Morphine on Pain-Related Brain Activation - An Experimental Functional Magnetic Resonance Imaging Study. Basic Clin Pharmacol Toxicol, 117(5), 316–322. doi:10.1111/bcpt.12415

Hansen, T. M., Olesen, A. E., Simonsen, C. W., Fischer, I. W., Lelic, D., Drewes, A. M., & Frokjaer, J. B. (2016). Acute Metabolic Changes Associated With Analgesic Drugs: An MR Spectroscopy Study. J Neuroimaging, 26(5), 545–551. doi:10.1111/jon.12345

Hautvast, R. W., Ter Horst, G. J., DeJong, B. M., DeJongste, M. J., Blanksma, P. K., Paans, A. M., & Korf, J. (1997). Relative changes in regional cerebral blood flow during spinal cord stimulation in patients with refractory angina pectoris. Eur J Neurosci, 9(6), 1178–1183. doi:10.1111/j.1460-9568.1997.tb01472.x

Holmes, C. J., Hoge, R., Collins, L., Woods, R., Toga, A. W., & Evans, A. C. (1998). Enhancement of MR images using registration for signal averaging. J Comput Assist Tomogr, 22(2), 324–333. doi:10.1097/00004728-199803000-00032

Hsieh, J. C., Hagermark, O., Stahle-Backdahl, M., Ericson, K., Eriksson, L., Stone-Elander, S., & Ingvar, M. (1994). Urge to scratch represented in the human cerebral cortex during itch. J Neurophysiol, 72(6), 3004–3008. doi:10.1152/jn.1994.72.6.3004

Huppert, T. J., Diamond, S. G., Franceschini, M. A., & Boas, D. A. (2009). HomER: a review of time-series analysis methods for near-infrared spectroscopy of the brain. Appl Opt, 48(10), D280–298. doi:10.1364/ao.48.00d280

Jeha, G. M., Kodumudi, V., O’Quinn, M. C., Luckett, K. O., Dickerson, T. G., Kaye, R. J., … Kaye, A. D. (2021). Management of Acute and Chronic Pain Associated With Hidradenitis Suppurativa: A Comprehensive Review of Pharmacologic and Therapeutic Considerations in Clinical Practice. Cutis, 108(5), 281–286. doi:10.12788/cutis.0383

Karunakaran, K. D., Peng, K., Berry, D., Green, S., Labadie, R., Kussman, B., & Borsook, D. (2021). NIRS measures in pain and analgesia: Fundamentals, features, and function. Neurosci Biobehav Rev, 120, 335–353. doi:10.1016/j.neubiorev.2020.10.023

Karunakaran, K. D., Peng, K., Green, S., Sieberg, C. B., Mizrahi-Arnaud, A., Gomez-Morad, A., … Borsook, D. (2023). Can pain under anesthesia be measured? Pain-related brain function using functional near-infrared spectroscopy during knee surgery. Neurophotonics, 10(2), 025014. doi:10.1117/1.NPh.10.2.025014

Kong, J., Loggia, M. L., Zyloney, C., Tu, P., LaViolette, P., & Gollub, R. L. (2010). Exploring the brain in pain: activations, deactivations and their relation. Pain, 148(2), 257–267. doi:10.1016/j.pain.2009.11.008

Krummenacher, P., Candia, V., Folkers, G., Schedlowski, M., & Schonbachler, G. (2010). Prefrontal cortex modulates placebo analgesia. Pain, 148(3), 368–374. doi:10.1016/j.pain.2009.09.033

Kucyi, A., Moayedi, M., Weissman-Fogel, I., Goldberg, M. B., Freeman, B. V., Tenenbaum, H. C., & Davis, K. D. (2014). Enhanced medial prefrontal-default mode network functional connectivity in chronic pain and its association with pain rumination. J Neurosci, 34(11), 3969–3975. doi:10.1523/JNEUROSCI.5055-13.2014

Kuijpers, T., van Middelkoop, M., Rubinstein, S. M., Ostelo, R., Verhagen, A., Koes, B. W., & van Tulder, M. W. (2011). A systematic review on the effectiveness of pharmacological interventions for chronic non-specific low-back pain. Eur Spine J, 20(1), 40–50. doi:10.1007/s00586-010-1541-4

Kussman, B. D., Aasted, C. M., Yucel, M. A., Steele, S. C., Alexander, M. E., Boas, D. A., … Becerra, L. (2016). Capturing Pain in the Cortex during General Anesthesia: Near Infrared Spectroscopy Measures in Patients Undergoing Catheter Ablation of Arrhythmias. PLoS One, 11(7), e0158975. doi:10.1371/journal.pone.0158975

Leknes, S., & Tracey, I. (2008). A common neurobiology for pain and pleasure. Nat Rev Neurosci, 9(4), 314–320. doi:10.1038/nrn2333

Lobanov, O. V., Quevedo, A. S., Hadsel, M. S., Kraft, R. A., & Coghill, R. C. (2013). Frontoparietal mechanisms supporting attention to location and intensity of painful stimuli. Pain, 154(9), 1758–1768. doi:10.1016/j.pain.2013.05.030

Lorenz, J., Minoshima, S., & Casey, K. L. (2003). Keeping pain out of mind: the role of the dorsolateral prefrontal cortex in pain modulation. Brain, 126(Pt 5), 1079–1091. doi:10.1093/brain/awg102

Lui, F., Colloca, L., Duzzi, D., Anchisi, D., Benedetti, F., & Porro, C. A. (2010). Neural bases of conditioned placebo analgesia. Pain, 151(3), 816–824. doi:10.1016/j.pain.2010.09.021

Lui, F., Duzzi, D., Corradini, M., Serafini, M., Baraldi, P., & Porro, C. A. (2008). Touch or pain? Spatio-temporal patterns of cortical fMRI activity following brief mechanical stimuli. Pain, 138(2), 362–374. doi:10.1016/j.pain.2008.01.010

Lundberg, S. M., & Lee, S.-I. (2017). A unified approach to interpreting model predictions. Paper presented at the Proceedings of the 31st International Conference on Neural Information Processing Systems, Long Beach, California, USA.

Mandrekar, J. N. (2010). Receiver operating characteristic curve in diagnostic test assessment. J Thorac Oncol, 5(9), 1315–1316. doi:10.1097/JTO.0b013e3181ec173d

Martucci, K. T., & Mackey, S. C. (2018). Neuroimaging of Pain: Human Evidence and Clinical Relevance of Central Nervous System Processes and Modulation. Anesthesiology, 128(6), 1241–1254. doi:10.1097/ALN.0000000000002137

Metz, C. E. (1978). Basic principles of ROC analysis. Semin Nucl Med, 8(4), 283–298. doi:10.1016/s0001-2998(78)80014-2

Molavi, B., & Dumont, G. A. (2012). Wavelet-based motion artifact removal for functional near-infrared spectroscopy. Physiol Meas, 33(2), 259–270. doi:10.1088/0967-3334/33/2/259

Montero-Hernandez, S., Orihuela-Espina, F., Sucar, E. L., Pinti, P., Hamilton, A., Burgess, P., & Tachtsidis, I. (2018). Estimating Functional Connectivity Symmetry between Oxy- and Deoxy-Haemoglobin: Implications for fNIRS Connectivity Analysis. Algorithms, 11(5). doi:10.3390/a11050070

Morton, D. L., Sandhu, J. S., & Jones, A. K. (2016). Brain imaging of pain: state of the art. J Pain Res, 9, 613–624. doi:10.2147/JPR.S60433

Okamoto, M., Dan, H., Sakamoto, K., Takeo, K., Shimizu, K., Kohno, S., … Dan, I. (2004). Three-dimensional probabilistic anatomical cranio-cerebral correlation via the international 10-20 system oriented for transcranial functional brain mapping. Neuroimage, 21(1), 99–111.

Ong, W. Y., Stohler, C. S., & Herr, D. R. (2019). Role of the Prefrontal Cortex in Pain Processing. Mol Neurobiol, 56(2), 1137–1166. doi:10.1007/s12035-018-1130-9

Ozturk, O., Algun, Z. C., Bombaci, H., & Erdogan, S. B. (2021). Changes in prefrontal cortex activation with exercise in knee osteoarthritis patients with chronic pain: An fNIRS study. J Clin Neurosci, 90, 144–151. doi:10.1016/j.jocn.2021.05.055

Paquette, T., Jeffrey-Gauthier, R., Leblond, H., & Pich, E. M. (2018). Functional Neuroimaging of Nociceptive and Pain-Related Activity in the Spinal Cord and Brain: Insights From Neurovascular Coupling Studies. Anat Rec (Hoboken), 301(9), 1585–1595. doi:10.1002/ar.23854

Pariente, J., White, P., Frackowiak, R. S., & Lewith, G. (2005). Expectancy and belief modulate the neuronal substrates of pain treated by acupuncture. Neuroimage, 25(4), 1161–1167. doi:10.1016/j.neuroimage.2005.01.016

Park, H. J., & Moon, D. E. (2010). Pharmacologic management of chronic pain. Korean J Pain, 23(2), 99–108. doi:10.3344/kjp.2010.23.2.99

Pedregosa, F., Varoquaux, G., Gramfort, A., Michel, V., Thirion, B., Grisel, O., … Duchesnay, É. (2011). Scikit-learn: Machine Learning in Python. J. Mach. Learn. Res., 12(null), 2825–2830.

Peng, K., Deepti Karunakaran, K., Lee, A., Gomez-Morad, A., Labadie, R., Mizrahi-Arnaud, A., … Borsook, D. (2021). Rhythmic Change of Cortical Hemodynamic Signals Associated with Ongoing Nociception in Awake and Anesthetized Individuals: An Exploratory Functional Near Infrared Spectroscopy Study. Anesthesiology, 135(5), 877–892. doi:10.1097/ALN.0000000000003986

Peng, K., Steele, S. C., Becerra, L., & Borsook, D. (2018). Brodmann area 10: Collating, integrating and high level processing of nociception and pain. Prog Neurobiol, 161, 1–22. doi:10.1016/j.pneurobio.2017.11.004

Peng, K., Yucel, M. A., Steele, S. C., Bittner, E. A., Aasted, C. M., Hoeft, M. A., … Borsook, D. (2018). Morphine Attenuates fNIRS Signal Associated With Painful Stimuli in the Medial Frontopolar Cortex (medial BA 10). Front Hum Neurosci, 12, 394. doi:10.3389/fnhum.2018.00394

Peterson, S. M., Steine-Hanson, Z., Davis, N., Rao, R. P. N., & Brunton, B. W. (2021). Generalized neural decoders for transfer learning across participants and recording modalities. J Neural Eng, 18(2). doi:10.1088/1741-2552/abda0b

Petrides, M., & Pandya, D. N. (2007). Efferent association pathways from the rostral prefrontal cortex in the macaque monkey. J Neurosci, 27(43), 11573–11586. doi:10.1523/JNEUROSCI.2419-07.2007

Petrovic, P., Kalso, E., Petersson, K. M., Andersson, J., Fransson, P., & Ingvar, M. (2010). A prefrontal non-opioid mechanism in placebo analgesia. Pain, 150(1), 59–65. doi:10.1016/j.pain.2010.03.011

Petrovic, P., Kalso, E., Petersson, K. M., & Ingvar, M. (2002). Placebo and opioid analgesia--imaging a shared neuronal network. Science, 295(5560), 1737–1740. doi:10.1126/science.1067176

Peyron, R., Laurent, B., & Garcia-Larrea, L. (2000). Functional imaging of brain responses to pain. A review and meta-analysis (2000). Neurophysiol Clin, 30(5), 263–288. doi:10.1016/s0987-7053(00)00227-6

Porro, C. A., Cettolo, V., Francescato, M. P., & Baraldi, P. (1998). Temporal and intensity coding of pain in human cortex. J Neurophysiol, 80(6), 3312–3320. doi:10.1152/jn.1998.80.6.3312

Racek, A. J., Hu, X., Nascimento, T. D., Bender, M. C., Khatib, L., Chiego, D., Jr., … DaSilva, A. F. (2015). Different Brain Responses to Pain and Its Expectation in the Dental Chair. J Dent Res, 94(7), 998–1003. doi:10.1177/0022034515581642

Rojas, R. F., Romero, J., Lopez-Aparicio, J., & Ou, K. L. (2021, 4-6 May 2021). Pain Assessment based on fNIRS using Bi-LSTM RNNs. Paper presented at the 2021 10th International IEEE/EMBS Conference on Neural Engineering (NER).

Seminowicz, D. A., & Davis, K. D. (2006). Cortical responses to pain in healthy individuals depends on pain catastrophizing. Pain, 120(3), 297–306. doi:10.1016/j.pain.2005.11.008

Seminowicz, D. A., & Moayedi, M. (2017). The Dorsolateral Prefrontal Cortex in Acute and Chronic Pain. J Pain, 18(9), 1027–1035. doi:10.1016/j.jpain.2017.03.008

Shapley, L. S. (1953). Contributions to the Theory of Games (AM-28), Volume II 17. A Value for n-Person Games. In H. W. Kuhn & A. W. Tucker (Eds.), (pp. 307–318): Princeton University Press.

Shrikumar, A., Greenside, P., & Kundaje, A. (2017). Learning important features through propagating activation differences. Paper presented at the Proceedings of the 34th International Conference on Machine Learning - Volume 70, Sydney, NSW, Australia.

Svensson, P., Minoshima, S., Beydoun, A., Morrow, T. J., & Casey, K. L. (1997). Cerebral processing of acute skin and muscle pain in humans. J Neurophysiol, 78(1), 450–460. doi:10.1152/jn.1997.78.1.450

Szucs, D., & Ioannidis, J. P. (2020). Sample size evolution in neuroimaging research: An evaluation of highly-cited studies (1990-2012) and of latest practices (2017-2018) in high-impact journals. Neuroimage, 221, 117164. doi:10.1016/j.neuroimage.2020.117164

Tanji, J. (1996). New concepts of the supplementary motor area. Curr Opin Neurobiol, 6(6), 782–787. doi:10.1016/s0959-4388(96)80028-6

Tetreault, P., Mansour, A., Vachon-Presseau, E., Schnitzer, T. J., Apkarian, A. V., & Baliki, M. N. (2016). Brain Connectivity Predicts Placebo Response across Chronic Pain Clinical Trials. PLoS Biol, 14(10), e1002570. doi:10.1371/journal.pbio.1002570

Tinnermann, A., Sprenger, C., & Buchel, C. (2022). Opioid analgesia alters corticospinal coupling along the descending pain system in healthy participants. Elife, 11. doi:10.7554/eLife.74293

Tseng, M. T., Tseng, W. Y., Chao, C. C., Lin, H. E., & Hsieh, S. T. (2010). Distinct and shared cerebral activations in processing innocuous versus noxious contact heat revealed by functional magnetic resonance imaging. Hum Brain Mapp, 31(5), 743–757. doi:10.1002/hbm.20902

Vachon-Presseau, E., Berger, S. E., Abdullah, T. B., Huang, L., Cecchi, G. A., Griffith, J. W., … Apkarian, A. V. (2018). Brain and psychological determinants of placebo pill response in chronic pain patients. Nat Commun, 9(1), 3397. doi:10.1038/s41467-018-05859-1

Vossel, S., Geng, J. J., & Fink, G. R. (2014). Dorsal and ventral attention systems: distinct neural circuits but collaborative roles. Neuroscientist, 20(2), 150–159. doi:10.1177/1073858413494269

Voytek, B., Davis, M., Yago, E., Barcelo, F., Vogel, E. K., & Knight, R. T. (2010). Dynamic neuroplasticity after human prefrontal cortex damage. Neuron, 68(3), 401–408. doi:10.1016/j.neuron.2010.09.018

Wager, T. D., Atlas, L. Y., Leotti, L. A., & Rilling, J. K. (2011). Predicting individual differences in placebo analgesia: contributions of brain activity during anticipation and pain experience. J Neurosci, 31(2), 439–452. doi:10.1523/JNEUROSCI.3420-10.2011

Wager, T. D., Atlas, L. Y., Lindquist, M. A., Roy, M., Woo, C. W., & Kross, E. (2013). An fMRI-based neurologic signature of physical pain. N Engl J Med, 368(15), 1388–1397. doi:10.1056/NEJMoa1204471

Wager, T. D., Rilling, J. K., Smith, E. E., Sokolik, A., Casey, K. L., Davidson, R. J., … Cohen, J. D. (2004). Placebo-induced changes in FMRI in the anticipation and experience of pain. Science, 303(5661), 1162–1167. doi:10.1126/science.1093065

Wanigasekera, V., Lee, M. C., Rogers, R., Kong, Y., Leknes, S., Andersson, J., & Tracey, I. (2012). Baseline reward circuitry activity and trait reward responsiveness predict expression of opioid analgesia in healthy subjects. Proc Natl Acad Sci U S A, 109(43), 17705–17710. doi:10.1073/pnas.1120201109

Watson, A., El-Deredy, W., Iannetti, G. D., Lloyd, D., Tracey, I., Vogt, B. A., … Jones, A. K. (2009). Placebo conditioning and placebo analgesia modulate a common brain network during pain anticipation and perception. Pain, 145(1-2), 24–30. doi:10.1016/j.pain.2009.04.003

Wen, Q., Sun, L., Yang, F., Song, X., Gao, J., Wang, X., & Xu, H. (2021). Time Series Data Augmentation for Deep Learning: A Survey. Paper presented at the Proceedings of the Thirtieth International Joint Conference on Artificial Intelligence, IJCAI-21. 10.24963/ijcai.2021/631 http://dx.doi.org/10.24963/ijcai.2021/631 https://www.ijcai.org/proceedings/2021/0631.pdf

Wiech, K., Kalisch, R., Weiskopf, N., Pleger, B., Stephan, K. E., & Dolan, R. J. (2006). Anterolateral prefrontal cortex mediates the analgesic effect of expected and perceived control over pain. J Neurosci, 26(44), 11501–11509. doi:10.1523/JNEUROSCI.2568-06.2006

Wise, S. P. (1985). The primate premotor cortex: past, present, and preparatory. Annu Rev Neurosci, 8, 1–19. doi:10.1146/annurev.ne.08.030185.000245

Woo, C. W., Chang, L. J., Lindquist, M. A., & Wager, T. D. (2017). Building better biomarkers: brain models in translational neuroimaging. Nat Neurosci, 20(3), 365–377. doi:10.1038/nn.4478

Wu, D., Jiang, X., & Peng, R. (2022). Transfer learning for motor imagery based brain– computer interfaces: A tutorial. Neural Networks, 153, 235–253. doi:10.1016/j.neunet.2022.06.008

Ye, J. C., Tak, S., Jang, K. E., Jung, J., & Jang, J. (2009). NIRS-SPM: statistical parametric mapping for near-infrared spectroscopy. Neuroimage, 44(2), 428–447. doi:10.1016/j.neuroimage.2008.08.036

Yucel, M. A., Aasted, C. M., Petkov, M. P., Borsook, D., Boas, D. A., & Becerra, L. (2015). Specificity of hemodynamic brain responses to painful stimuli: a functional near-infrared spectroscopy study. Sci Rep, 5, 9469. doi:10.1038/srep09469

Yucel, M. A., Selb, J., Aasted, C. M., Lin, P. Y., Borsook, D., Becerra, L., & Boas, D. A. (2016). Mayer waves reduce the accuracy of estimated hemodynamic response functions in functional near-infrared spectroscopy. Biomed Opt Express, 7(8), 3078–3088. doi:10.1364/BOE.7.003078

Zhang, Y., Brooks, D. H., Franceschini, M. A., & Boas, D. A. (2005). Eigenvector-based spatial filtering for reduction of physiological interference in diffuse optical imaging. J Biomed Opt, 10(1), 11014. doi:10.1117/1.1852552

Zunhammer, M., Spisak, T., Wager, T. D., Bingel, U., & Placebo Imaging, C. (2021). Metaanalysis of neural systems underlying placebo analgesia from individual participant fMRI data. Nat Commun, 12(1), 1391. doi:10.1038/s41467-021-21179-3

