## Supplementary material for "Explainable fNIRS Based Pain Decoding Under Pharmacological Conditions via Deep Transfer Learning Approach": Highlights

•	Pain decoding using transfer learning (TL) was performed under 6 distinct analgesic conditions.

•	Knowledge transfer from a pre-drug base model resulted in highly accurate post-drug pain decoding.

•	Post-placebo models had higher decoding accuracy than post-morhine models.

•	DeepSHAP algorithm was adapted to decoding models to predict cortical contribution weights.

•	Contribution of different ROIs to classification performance changes under analgesic states.

- Unveiling the explanation power of different ROIs might aid the design of efficient fNIRS-BCIs.
