## Supplementary material for "Explainable fNIRS Based Pain Decoding Under Pharmacological Conditions via Deep Transfer Learning Approach": Data and Code Availability

In addition to HomER3, Tensorflow toolkit (version 2.8.0) and Shap toolbox, the code can be downloaded from the website <https://github.com/aykuteken/Pain_decoding/> The data information can be found in <https://www.nitrc.org/projects/yucel18pain/>.
