## Supplementary material for "Explainable fNIRS Based Pain Decoding Under Pharmacological Conditions via Deep Transfer Learning Approach": Table1

|  | Pre Drug Session | | | | | Morphine Session | | | | | Placebo Session | | | | |
| --- | --- | --- | --- | --- | --- | --- | --- | --- | --- | --- | --- | --- | --- | --- | --- |
| Channel  Number | Mean X | Mean Y | Mean Z | Std. Dev. | **Corr. Reg.** | Mean X | Mean Y | Mean Z | Std. Dev. | **Corr. Reg.** | Mean X | Mean Y | Mean Z | Std. Dev. | **Corr. Reg.** |
| 1 | -63.45 | 4.05 | 25.05 | 5.25 | **L PMC** | -63.58 | 3.69 | 24.58 | 5.34 | **L PMC** | -63.31 | 4.41 | 25.51 | 5.16 | **L PMC** |
| 2 | -53.77 | 10.19 | 43.02 | 5.50 | **L DLPFC** | -54.11 | 10.53 | 42.11 | 5.65 | **L DLPFC** | -53.44 | 9.85 | 43.92 | 5.35 | **L PMC** |
| 3 | -59.52 | 17.76 | 19.86 | 5.15 | **L IFG** | -59.75 | 17.17 | 19.11 | 5.17 | **L IFG** | -59.28 | 18.36 | 20.62 | 5.13 | **L IFG** |
| 4 | -56.50 | 29.08 | 14.62 | 5.78 | **L IFG** | -57.00 | 28.39 | 13.75 | 5.73 | **L IFG** | -56.00 | 29.77 | 15.49 | 5.83 | **L IFG** |
| 5 | -50.90 | 23.54 | 38.03 | 5.40 | **L DLPFC** | -51.31 | 23.75 | 37.00 | 5.38 | **L DLPFC** | -50.49 | 23.33 | 39.05 | 5.41 | **L DLPFC** |
| 6 | -47.47 | 33.66 | 34.12 | 5.62 | **L DLPFC** | -48.19 | 33.61 | 32.83 | 5.75 | **L DLPFC** | -46.74 | 33.72 | 35.41 | 5.48 | **L DLPFC** |
| 7 | -52.44 | 40.15 | 9.17 | 5.91 | **L DLPFC** | -53.03 | 39.22 | 8.19 | 5.77 | **L DLPFC** | -51.85 | 41.08 | 10.15 | 6.04 | **L DLPFC** |
| 8 | -42.99 | 44.76 | 28.61 | 5.79 | **L DLPFC** | -43.83 | 44.53 | 27.39 | 5.85 | **L DLPFC** | -42.15 | 45.00 | 29.82 | 5.72 | **L DLPFC** |
| 9 | -19.56 | 69.51 | 14.11 | 5.81 | **L FPA** | -19.94 | 69.67 | 13.17 | 5.44 | **L FPA** | -19.18 | 69.36 | 15.05 | 6.17 | **L FPA** |
| 10 | -17.47 | 58.52 | 35.89 | 5.43 | **L DLPFC** | -17.81 | 58.92 | 35.11 | 5.42 | **L DLPFC** | -17.13 | 58.13 | 36.67 | 5.43 | **L DLPFC** |
| 11 | -6.25 | 70.57 | 13.36 | 6.24 | **L FPA** | -6.75 | 71.19 | 12.61 | 5.89 | **L FPA** | -5.74 | 69.95 | 14.10 | 6.59 | **L FPA** |
| 12 | 9.98 | 70.96 | 14.35 | 6.61 | **R FPA** | 8.58 | 71.19 | 13.53 | 6.45 | **R FPA** | 11.39 | 70.72 | 15.18 | 6.77 | **R FPA** |
| 13 | -5.52 | 61.40 | 35.19 | 5.78 | **L DLPFC** | -5.83 | 62.00 | 34.58 | 5.64 | **L FPA** | -5.21 | 60.80 | 35.80 | 5.91 | **L DLPFC** |
| 14 | 7.95 | 60.90 | 35.92 | 6.33 | **R DLPFC** | 6.36 | 61.50 | 35.17 | 6.21 | **R DLPFC** | 9.54 | 60.31 | 36.67 | 6.45 | **R DLPFC** |
| 15 | 24.39 | 68.41 | 15.18 | 6.45 | **R FPA** | 22.78 | 69.25 | 14.97 | 5.92 | **R FPA** | 26.00 | 67.56 | 15.39 | 6.98 | **R FPA** |
| 16 | 21.01 | 57.21 | 37.25 | 6.02 | **R DLPFC** | 19.50 | 58.17 | 37.06 | 5.32 | **R DLPFC** | 22.51 | 56.26 | 37.44 | 6.71 | **R DLPFC** |
| 17 | 36.39 | -8.49 | 68.76 | 5.85 | **R PMC** | 34.86 | -6.61 | 69.17 | 5.25 | **R PMC** | 37.92 | -10.36 | 68.36 | 6.45 | **R PMC** |
| 18 | 38.04 | -29.90 | 71.17 | 5.26 | **R MI** | 36.78 | -27.75 | 72.11 | 4.24 | **R MI** | 39.31 | -32.05 | 70.23 | 6.28 | **R SI** |
| 19 | 45.37 | -8.26 | 63.41 | 4.88 | **R PMC** | 44.72 | -6.53 | 63.22 | 4.84 | **R PMC** | 46.03 | -10.00 | 63.59 | 4.92 | **R PMC** |
| 20 | 54.51 | -8.50 | 55.52 | 5.03 | **R PMC** | 53.81 | -6.56 | 55.78 | 4.61 | **R PMC** | 55.21 | -10.44 | 55.26 | 5.44 | **R PMC** |
| 21 | 46.96 | -29.95 | 65.71 | 4.30 | **R SI** | 46.69 | -28.14 | 66.19 | 4.07 | **R SI** | 47.23 | -31.77 | 65.23 | 4.53 | **R SI** |
| 22 | 56.01 | -31.63 | 57.72 | 4.64 | **R SMG** | 55.56 | -29.64 | 58.28 | 4.29 | **R SMG** | 56.46 | -33.62 | 57.15 | 4.99 | **R SMG** |
| 23 | 61.34 | -9.85 | 46.81 | 5.49 | **R PMC** | 61.11 | -8.17 | 46.64 | 4.70 | **R PMC** | 61.56 | -11.54 | 46.98 | 6.27 | **R PMC** |
| 24 | 63.14 | -33.60 | 49.70 | 5.20 | **R SMG** | 63.61 | -31.94 | 49.61 | 4.37 | **R SMG** | 62.67 | -35.26 | 49.80 | 6.02 | **R SMG** |

Table 1. Mean MNI coordinates of long channels averaged across all subjects and scans for pre-drug, morphine and placebo administration sessions. The corresponding anatomical regions are given in Talairach Space. Std. Dev. : Standard Deviation, Corr. Reg. : Corresponding Region, L: Left, R: Right, PMC: Pre-motor cortex, IFG: Inferior Frontal Gyrus, SI : Primary Somatosensory Cortex, MI: Primary Motor Cortex, SMG: Supramarginal Gyrus, FPA: Frontopolar area, DLPFC: Dorsolateral prefrontal cortex.
