## Supplementary material for "Explainable fNIRS Based Pain Decoding Under Pharmacological Conditions via Deep Transfer Learning Approach": Table2

Table 2. Test performances of PDM and all post-drug models in terms of their accuracy, sensitivity and specificity results averaged across 30 runs.

| ***Session*** | | ***Accuracy (Mean Std. Dev.)*** | ***Sensitivity (Mean Std. Dev.)*** | ***Specificity (Mean Std. Dev.)*** | ***AUC (Mean Std. Dev.)*** |
| --- | --- | --- | --- | --- | --- |
| *Pre-Drug* | | *0.97 ± 0.03* | *0.97± 0.04* | *0.97 0.04* | *0.97 0.03* |
| *Morphine* | 30 min | *0.91 0.05* | *0.90 0.08* | *0.91 0.05* | *0.91 0.05* |
|  | 60 min | *0.90 0.07* | *0.88 0.11* | *0.90 0.07* | *0.90 0.07* |
|  | 90 min | *0.91 0.05* | *0.89 0.08* | *0.91 0.05* | *0.91 0.05* |
| *Placebo* | 30 min | *0.92 0.06* | *0.92 0.08* | *0.92 0.06* | *0.92 0.06* |
|  | 60 min | *0.92 0.05* | *0.91 0.08* | *0.92 0.05* | *0.92 0.05* |
|  | 90 min | *0.91 0.07* | *0.91 0.08* | *0.91 0.07* | *0.91 0.07* |
